# NIMETOX-informed Precision Nomothetic Models of Major Depressive Disorder: Group, Phenome, and Individual Signatures

**DOI:** 10.64898/2026.01.23.26344678

**Authors:** Michael Maes, Mengqi Niu, Ping Wang, Annabel Maes, Yiping Luo, Chenkai Yangyang, Xiaoman Zhuang, Abbas F. Almulla, Jing Li, Yingqian Zhang

## Abstract

**Background:** Major depressive disorder (MDD) is a neuro-immune-metabolic-oxidative (NIMETOX) disorder. Nevertheless, the effects of alterations in immune responsiveness, oxidative stress, antioxidant defenses, gut-derived short-chain fatty acids (SCFAs), metabolic hormones and adipokines on metabolomic modules and the MDD phenome have remained elusive.

**Methods:** Serum samples from 125 MDD inpatients and 40 healthy controls were analyzed using high-resolution metabolomics assays (liquid chromatography, mass spectrometry) in conjunction with assays of 68 additional NIMETOX markers. A machine learning pipeline was implemented to delineate the associations between MDD, clinical phenome features, metabolomic modules, and 68 NIMETOX biomarkers.

**Results:** The metabolomics and NIMETOX biomarkers distinguished MDD from controls with a cross-validated accuracy of >95%. Core biomarkers of MDD encompass (in order of decreasing importance) diacylglycerol lipotoxicity, phospholipid remodeling, fatty acid signaling, mitochondrial-redox dysfunction, diminished antioxidant defenses (including decreased paraoxonase 1 activity, Apolipoprotein A1, reverse cholesterol transport, ether lipids), inflammatory response, increased epidermal growth factor, disbalances in gut-derived SCFAs, increased oxidized high-density lipoprotein cholesterol, and changes in metabolic hormones. A large part of the variance in overall severity of illness (76.3%), physiosomatic symptoms (61.9%), current suicidal ideation (40.6%), and recurrence of illness (28.8%) was explained by those pathways. Lipotoxicity, phospholipid remodeling, fatty acid storage, and clinical phenome features converge onto a singular latent construct—the metabolic phenome of MDD. The various NIMETOX pathways mediate the impact of adverse childhood experiences on metabolomics and MDD.

**Conclusions:** MDD is a NIMETOX disorder in which metabolomic signals represent a final common pathway underlying symptom severity, recurrence of illness, and suicidality.

## Introduction

The neuroimmune–metabolic–oxidative stress (NIMETOX) theory of major depressive disorder (MDD) posits that depression is driven by a convergence of neuroimmune activation, metabolic dysregulation, and oxidative–nitrosative toxicity, rather than isolated biomarker or neurotransmitter deficits (Maes, Almulla et al. 2025, Maes, Jirakran et al. 2025). In this framework, peripheral immune activation triggers sustained production of reactive oxygen and nitrogen species (ROS/RNS), leading to lipid, protein, and mitochondrial damage (Maes, Almulla et al. 2025, Maes, Jirakran et al. 2025). These processes disrupt membrane integrity, energy metabolism, and redox homeostasis, thereby impairing neuronal plasticity and survival. Altered lipid signaling and oxidative injury further amplify inflammatory pathways, creating a self-perpetuating cycle of toxicity. Through humoral, endothelial, and neural routes, these peripheral disturbances signal to the brain, inducing limbic–cortical circuit dysfunction (Maes, Almulla et al. 2025, Maes, Jirakran et al. 2025). NIMETOX thus provides a systems-biology model linking immune, metabolic, and oxidative pathways to the clinical heterogeneity and recurrence of MDD (Maes, Almulla et al. 2025, Maes, Almulla et al. 2025, Maes, Jirakran et al. 2025).

Building on the NIMETOX framework, Maes and colleagues have applied this model to develop precision–nomothetic approaches to depression, integrating NIMETOX biomarkers into coherent explanatory systems (Maes 2022, Maes and Stoyanov 2022, Maes, Moraes et al. 2023, Maes, Zhou et al. 2024). By quantifying multiple NIMETOX components simultaneously, these models move beyond group averages to identify biologically defined subgroups, such as those stratified by recurrence of illness (ROI) (Maes, Almulla et al. 2025). Importantly, NIMETOX variables can also be combined into individual-level composite scores, allowing the characterization of each patient’s unique pathophysiological profile (Maes and Almulla 2023, Maes, Moraes et al. 2023). This approach supports personalized treatment selection, as different patients exhibit distinct dominance of immune, redox, or metabolic pathways. Thus, NIMETOX provides a scalable framework spanning group-level inference, subgroup stratification, and individualized precision medicine in MDD (Maes and Stoyanov 2022).

Recently, it was shown that using untargeted serum metabolomics - a panel of 16 metabolites was disclosed (**Table 1**), which robustly discriminated MDD inpatients from healthy controls, achieving a near-perfect classification accuracy across multivariate models. These metabolites converged into six biologically coherent functional modules (**Table 1**), encompassing diacylglycerol (DAG) lipotoxicity, membrane phospholipid (PL) remodeling, aberrations in fatty acid storage/metabolism, mitochondrial redox overload, ether-lipid depletion, and retinoid deficiency, all of which are integral to the NIMETOX model of depression (Maes, Niu et al. 2025). Importantly, module-based analyses revealed marked heterogeneity within MDD, with individual patients displaying distinct dominance patterns across the six modules (Maes, Niu et al. 2025). These findings support a precision-medicine interpretation, whereby shared disease biology coexists with individualized pathway activation profiles that may inform tailored therapeutic strategies.

**Table 1.**
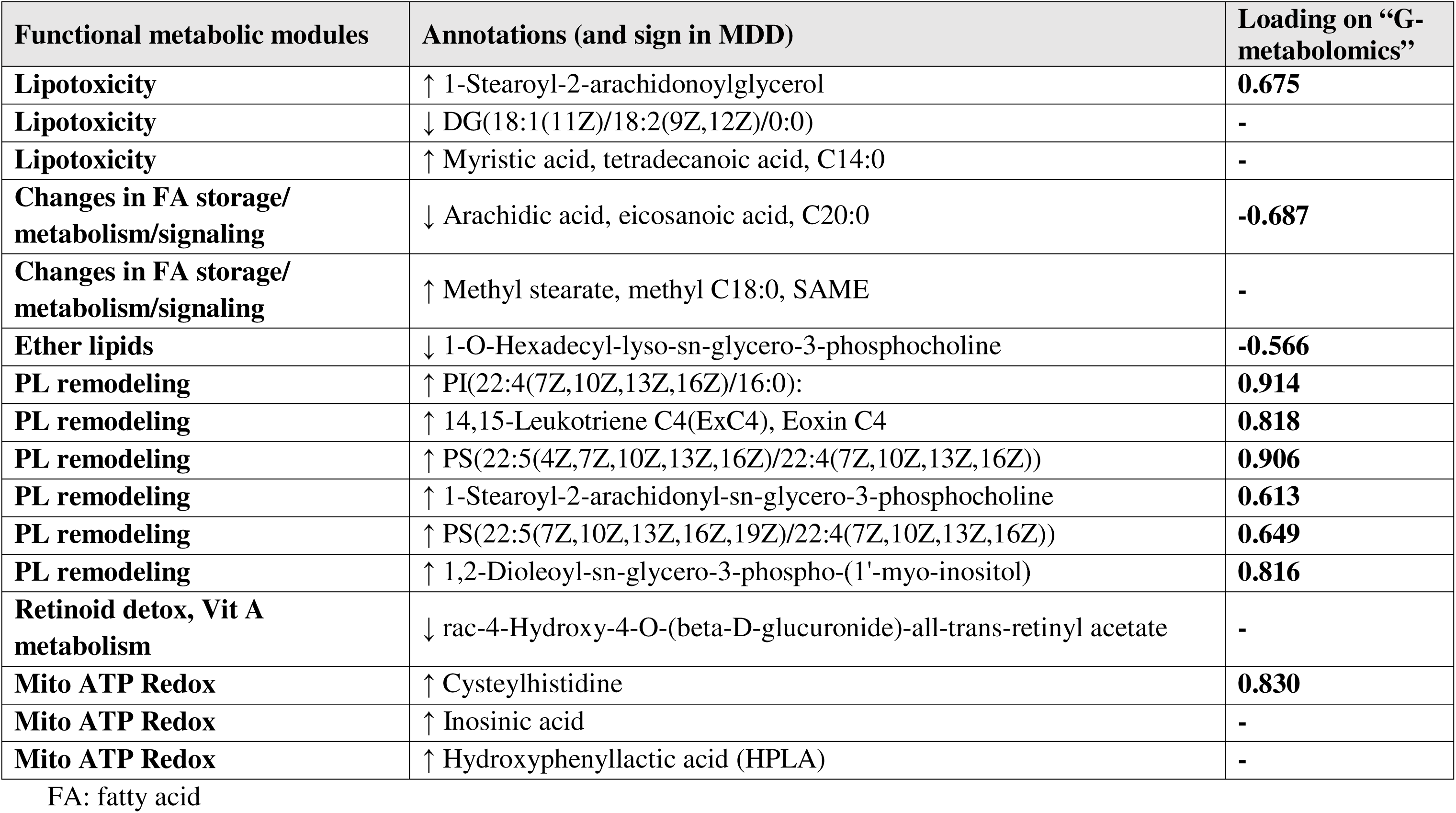
The metabolic modules examined in the present study.

| Functional metabolic modules | Annotations (and sign in MDD) | Loading on “G-metabolomics” |
| --- | --- | --- |
| **Lipotoxicity** | ↑ 1-Stearoyl-2-arachidonoylglycerol | **0.675** |
| **Lipotoxicity** | ↓ DG(18:1(11Z)/18:2(9Z,12Z)/0:0) | - |
| **Lipotoxicity** | ↑ Myristic acid, tetradecanoic acid, C14:0 | - |
| **Changes in FA storage/metabolism/signaling** | ↓ Arachidic acid, eicosanoic acid, C20:0 | **-0.687** |
| **Changes in FA storage/metabolism/signaling** | ↑ Methyl stearate, methyl C18:0, SAME | - |
| **Ether lipids** | ↓ 1-O-Hexadecyl-lyso-sn-glycero-3-phosphocholine | **-0.566** |
| **PL remodeling** | ↑ PI(22:4(7Z,10Z,13Z,16Z)/16:0): | **0.914** |
| **PL remodeling** | ↑ 14,15-Leukotriene C4(ExC4), Eoxin C4 | **0.818** |
| **PL remodeling** | ↑ PS(22:5(4Z,7Z,10Z,13Z,16Z)/22:4(7Z,10Z,13Z,16Z)) | **0.906** |
| **PL remodeling** | ↑ 1-Stearoyl-2-arachidonoyl-sn-glycero-3-phosphocholine | **0.613** |
| **PL remodeling** | ↑ PS(22:5(7Z,10Z,13Z,16Z,19Z)/22:4(7Z,10Z,13Z,16Z)) | **0.649** |
| **PL remodeling** | ↑ 1,2-Dioleoyl-sn-glycero-3-phospho-(1'-myo-inositol) | **0.816** |
| **Retinoid detox, Vit A metabolism** | ↓ rac-4-Hydroxy-4-O-(beta-D-glucuronide)-all-trans-retinyl acetate | - |
| **Mito ATP Redox** | ↑ Cysteylhistidine | **0.830** |
| **Mito ATP Redox** | ↑ Inosinic acid | - |
| **Mito ATP Redox** | ↑ Hydroxyphenyllactic acid (HPLA) | - |

FA: fatty acid

Within the NIMETOX framework, the metabolomic alterations observed in MDD are plausibly downstream consequences of converging NIMETOX abnormalities (Maes, Almulla et al. 2025). An activated acute phase response, marked by reduced albumin and transferrin, lowers antioxidant buffering and increases labile iron, thereby amplifying lipid peroxidation and membrane damage (Maes, Vandewoude et al. 1991, Maes 1993, Maes, Christophe et al. 1999, Maes, Almulla et al. 2025). Concurrent activation of the immune-inflammatory response system (IRS)—with enhanced T helper (Th)-1, M1 macrophage, and Th17 activity—drives cytokine signaling, notably tumor necrosis factor (TNF)-α, that stimulates PLA activation, arachidonic acid/adrenic phospholipid remodeling, and lipoxygenase (LOX)-derived eicosanoids (Maes, Jirakran et al. 2025). Elevated epidermal growth factor (EGF) and soluble CD40L (sCD40L) further potentiate immune–endothelial activation and intracellular signaling cascades (Niu, Zhang et al. 2025). Dysregulation of the compensatory immunoregulatory system (CIRS) may be insufficient to counterbalance these proinflammatory forces (Niu, Zhang et al. 2025). In parallel, reduced antioxidant defenses (enzymatic and non-enzymatic) and increased ROS/RNS exacerbate lipid, protein, and mitochondrial injury (Morris, Gevezova et al. 2022). Impairments in Apolipoprotein (Apo)A1, high-density lipoprotein cholesterol (HDL), paraoxonase (PON)1 and lecithin-cholestrol-acyltransferase (LCAT) activity, leading to reduced reverse cholesterol transport (RCT), might diminish lipid detoxification and antioxidative capacity (Maes, Zhou et al. 2024, Maes, Jirakran et al. 2025).

Higher short-chain fatty acids (SCFAs), such as acetate (AA), propionate (PA), butyrate (BA), are putatively protective across the metabolic modules by dampening TNF-driven inflammation, reducing eicosanoid flux, and lowering downstream DAG lipotoxic signaling via improved metabolic control (Koh, De Vadder et al. 2016, Dalile, Van Oudenhove et al. 2019). Butyrate is typically most potent, supporting barrier integrity, reducing oxidative burden, and indirectly preserving plasmalogens/ether lipids and limiting mitochondrial–redox stress (Koh, De Vadder et al. 2016, Ríos-Covián, Ruas-Madiedo et al. 2016, Dalile, Van Oudenhove et al. 2019). Elevated 2-methylbutyric acid (MBA) is generally interpreted as a marker of proteolytic fermentation of branched-chain amino acids, which often accompanies dysbiosis and mucosal stress (Morrison and Preston 2016, Rios-Covian, González et al. 2020).

Lower adiponectin can plausibly worsen the metabolic modules by removing an anti-inflammatory “brake” on TNF signaling, thereby facilitating PL remodeling and downstream DAG lipotoxicity, while also weakening mitochondrial resilience and antioxidant tone (Basati, Pourfarzam et al. 2011, Villarreal-Molina and Antuna-Puente 2012). Lower PAI-1 could indicate altered coagulation–endothelial setpoints; in a lipid–redox context it may accompany immune-metabolic shifts that still promote eicosanoid/oxidized phospholipid stress (Agren, Wiman et al. 2006, Fu and Birukov 2009).

Adverse childhood experiences (ACEs) exert long-lasting effects on NIMETOX homeostasis and are now recognized as upstream drivers of biological vulnerability in MDD (Maes, Almulla et al. 2025). ACEs are consistently associated with persistent IRS activation (Th1/Th17, M1 macrophage polarization), elevated proinflammatory cytokines (e.g., TNF-α, IL-6), and compensatory but insufficient CIRS responses (Maes, Rachayon et al. 2022, Maes, Almulla et al. 2025). These immune alterations promote oxidative and nitrosative stress, leading to depletion of endogenous antioxidants and increased lipid peroxidation (Moraes, Maes et al. 2018, Maes, Almulla et al. 2025). In parallel, ACEs are linked to atherogenic lipid profiles, reduced HDL/PON1 activity, impaired RCT, and changes in the gut microbiome (Maes, Zhou et al. 2024, Maes, Almulla et al. 2025). Through these mechanisms, ACEs can indirectly shape the lipid–redox metabolomic modules identified in MDD.

Despite extensive evidence for NIMETOX abnormalities in MDD, their integrated effects on metabolomic modules remain unknown. In particular, how established NIMETOX biomarkers jointly shape lipid–redox metabolic domains has not been systematically examined. Hence, the aims of this study are to (i) delineate the impact of changes in immune responsivity (cytokines/IRS/CIRS/EGF/sCD40L/acute phase response), oxidative stress, HDL–PON1/RCT, gut-microbiome-derived SCFAs, and aberration in metabolic hormones and adipokines on the metabolic modules in MDD; (ii) identify a set of NIMETOX variables that discriminate MDD from healthy controls, (iii) build nomothetic NIMETOX - metabolomic models of MDD, and (iv) explore the potential utility of the combined biomarker set for precision medicine, including individualized NIMETOX profiling.

## Methods

### Participants

This case–control, cross-sectional study involved 165 participants, consisting of 125 inpatients with acute severe MDD and 40 healthy controls, conducted at the International NIMETOX Center, Mental Health Center of Sichuan Provincial People’s Hospital, Chengdu, China. Participants ranged in age from 18 to 65 years, with an equitable sex distribution. Inpatients with MDD were diagnosed based on DSM-5 criteria; they were evaluated during an acute episode of severe MDD and exhibited a Hamilton Depression Rating Scale-21 (HAMD-21) score exceeding 18 (Hamilton 1960, Niu, Zhang et al. 2025). The control group was composed of hospital workers, their family, and acquaintances of patients, and was group- or frequency-matched to the MDD group based on age, sex, education level, and body mass index (BMI). Participants in the control group were excluded if they had a current or lifetime diagnosis of MDD, dysthymia, any DSM-5 anxiety disorder, or a family history of mood disorders, substance use disorders (except nicotine dependence), or suicide. All participants, or their legal representatives when applicable, furnished written informed consent before inclusion. The Ethics Committee of Sichuan Provincial People’s Hospital authorized the study protocol (Ethics [Research] No. 2024-203).

Participants were excluded if they met any of the following criteria: (i) current or lifetime diagnosis of major psychiatric disorders other than MDD, including schizophrenia spectrum disorders, bipolar disorder, schizoaffective disorder, eating disorders, substance use disorders (except nicotine dependence), psycho-organic disorders, autism spectrum disorder, or antisocial and borderline personality disorders; (ii) presence of neurological illnesses such as epilepsy, stroke, multiple sclerosis, Alzheimer’s disease, Parkinson’s disease, or brain tumors; (iii) active or recent (within the past three months) infectious disease, or a history of severe allergic reactions within the preceding month; (iv) chronic systemic or inflammatory medical conditions, including rheumatoid arthritis, inflammatory bowel disease, type 1 diabetes, chronic obstructive pulmonary disease, systemic lupus erythematosus, psoriasis, or malignancy; (v) current use of immunosuppressive agents, corticosteroids, or other medications known to substantially affect immune function; (vi) pregnancy or breastfeeding; (vii) major surgery within the previous three months; (viii) frequent use of analgesic medications; or (ix) use of therapeutic doses of antioxidant supplements or omega-3 fatty acids during the three months prior to enrollment.

### Clinical Assessment

Semi-structured interviews were conducted by a qualified physician to gather medical, psychiatric, and family histories in addition to demographic data. The Mini International Neuropsychiatric Interview (M.I.N.I.), which evaluates both present and past major DSM-5 psychiatric illnesses, was used to confirm mental diagnoses. As previously mentioned, participants filled out a series of standardized questionnaires on the same day to gauge their level of anxiety, depression, and somatic-psychosomatic symptoms (Niu, Zhang et al. 2025).

The HAMD-21 was used to measure depression severity (Hamilton 1960), and the Beck Depression Inventory-II (BDI-II) (Beck, Steer et al. 1996) was used to measure self-reported depression. The Hamilton Anxiety Rating Scale (HAMA) was used to measure anxiety severity, and the State-Trait Anxiety Inventory state (STAI) subscale was used to measure self-reported state anxiety (Hamilton 1959, Hamilton 1960, Spielberger, Gorsuch et al. 1983, Beck, Steer et al. 1996). The severity of symptoms associated with chronic fatigue syndrome (CFS) was evaluated using the 12-item FibroFatigue (FF) Scale, a clinical interview instrument (Zachrisson, Regland et al. 2002). The severity of somatic symptoms that people had over the previous seven days was measured using the Somatic Symptom Scale-8 (SSS-8) (Gierk, Kohlmann et al. 2014). Suicidal ideation (SI) and lifetime and current suicidal attempts (SA) were evaluated using the Columbia Suicide Severity Rating Scale (C-SSRS) (Posner, Brown et al. 2011). As previously mentioned, overall severity of disorder (OSOD), physiosomatic symptoms, and current suicidal ideation scores were calculated using these scales (Niu, Zhang et al. 2025). The recurrence of illness (ROI) index was calculated in this study using two items from the C-SSRS: the number of lifetime suicidal attempts and ideation prior to the index episode. The z-scores for the number of depressive episodes, suicide attempts, and suicidal thoughts across a person’s lifetime are added to create the ROI, a z unit-based composite score (Maes, Zhou et al. 2024, Maes, Almulla et al. 2025). A validated Chinese version of the Childhood Trauma Questionnaire Short Form (CTQ-SF) (Bernstein, Stein et al. 2003) was used to assess ACEs (Zhao, Zhang et al. 2005). Emotional abuse, physical abuse, sexual abuse, emotional neglect, and physical neglect were the five subscales for which scores were calculated (Niu, Zhang et al. 2025).

The components of metabolic syndrome (MetS), including height, weight, BMI, and waist circumference, were examined in this study. Weight in kilograms was divided by height in meters squared to determine BMI. The measurement of waist circumference was taken halfway between the inferior rib and the iliac crest. The 2009 joint statement from the National Heart, Lung, and Blood Institute and the American Heart Association defined MetS (Alberti, Eckel et al. 2009). Three or more of the following five criteria were met to diagnosis MetS: i) Triglycerides ≥150 mg/dL; ii) male waist circumference ≥90 cm or female waist circumference ≥80 cm; iii) systolic blood pressure ≥130 mm Hg or diastolic blood pressure ≥85 mm Hg, or the use of antihypertensive drugs; iv) fasting glucose ≥100 mg/dL or diabetes mellitus; v) male HDL cholesterol <40 mg/dL or female HDL cholesterol <50 mg/dL.

### Assays

A disposable syringe was used to extract 30 mL of fasting venous blood, which was then transferred into serum tubes. Blood samples were collected between 06:30 and 8:00 a.m. Serum was collected, aliquoted into Eppendorf tubes, and kept at -80°C for subsequent analysis after centrifugation at 3500 rpm. The assay techniques for the various biomarkers (n=51 and 17 derived indices) were described in our earlier studies (Almulla, Niu et al. 2025, Almulla, Niu et al. 2025, Maes, Niu et al. 2025, Niu, Zhang et al. 2025) and are summarized in Electronic Supplementary File (ESF), Table 1. All samples for metabolomics experiments were analyzed using mass spectrometry (MS) and liquid chromatography (LC) techniques, as previously mentioned (Maes, Niu et al. 2025). A UPLC system (Vanquish, Thermo Scientific) connected to an electrospray ionization quadrupole-Orbitrap hybrid high-resolution mass spectrometer (Q Exactive HF-X, Thermo Scientific) was used for the LC-MS analysis. Three main processing steps were applied to metabolite data: first, data filtration to remove samples with more than 80% missing values or quality control (QC) samples with more than 50% missing data; second, data imputation using the K-nearest neighbor (KNN) method; and third, data standardization using Probabilistic Quotient Normalization (PQN). The process for determining metabolomics is described in ESF, Assays.

### Statistics

#### Classical statistical tests

Correlations between continuous data were analyzed using Pearson’s correlation coefficients. An analysis of variance was utilized to assess the differences across the various study groups for continuous variables, including demographic and clinical data. Contingency table analysis was employed to investigate relationships among variables using categorical data, utilizing Chi-square testing as the methodological approach. Multiple comparisons were subjected to False Discovery Rate (FDR) p correction. The construction of z unit-based composites and principal component analysis (PCA) were applied for feature reduction. A principal component was considered valid and interpretable as an underlying construct when the Kaiser–Meyer–Olkin (KMO) measure exceeded 0.70, the component explained more than 50% of the variance, and all variable loadings were greater than 0.55. The sample size estimate was derived from the findings of Maes et al. (Maes, Niu et al. 2025), which indicated that a significant portion of the variance in OSOD, i.e., more than 25% could be anticipated using multivariable regression analysis including metabolic or NIMETOX variables. Power calculation for the primary statistical analysis of this study, specifically multiple regression analyses examining the predictors of the phenome scores, was conducted using G*Power 3.1.9.4 software. The analysis utilized an effect size of 0.33 (approximately 25% of the phenome explained), an alpha level of 0.05 (two-tailed), a power of 0.8, and a maximum of seven covariates. The power analysis indicated a minimum necessary sample size of 51 based on the determined effect size. The sample size was further expanded to enhance the accuracy and stability of regression coefficient estimations, as well as to enable independent testing samples, even though a priori power analysis showed sufficient power (0.80).

#### Machine learning

Biomarker discovery was performed using a supervised multistage pipeline that included linear discriminant analysis (LDA), multiple regression analysis, CATREG lasso and ridge regression, principal component analysis (PCA), partial least squares discriminant analysis (PLS-DA), orthogonal PLS-DA (OPLS-DA), PLS-regression, PLS-structural equation modeling (PLS-SEM), support vector machine (SVM), and random forest. Furthermore, we utilized a rigorous sample splitting approach to avert model overfitting or circularity. Statistical analysis was mainly conducted using R (v4.0), Statistica 14.0, SPSS 30.0, SmartPLS, and the Unscrambler. Cluster heatmaps were generated utilizing the R program heatmap. PCA for significant differential metabolite analysis was performed using the R package metaX. PLS-DA was performed utilizing a) the R package ropls, subsequently calculating variable importance in projection (VIP) values for each variable; and b) Statistica 14.0, followed by the computation of variable importances and case-wise score contributions. Heatmaps, PLS-DA, and OPLS-DA figures were created via the OmicStudio program available at https://www.omicstudio.cn/tool. Centered Log-Ratio (CLR) transformations of the metabolic data were applied in R due to the compositional characteristics of metabolomics data, which is constrained by constant-sum scaling, rendering it susceptible to spurious correlations and deceptive patterns when assessed using raw or non-ratio-based scales.

#### Relationships between metabolomic modules and other biomarkers

The relationships between the metabolic modules and other NIMETOX biomarkers were visualized using heatmaps based on Pearson’s correlation coefficients. In addition, we performed multiple regression analyses (including ridge, lasso, and elastic-net regression) to determine the most important biomarkers predicting the metabolic modules established previously. Alongside the manual multiple regression method, we employed automated regression techniques to ascertain the NIMETOX biomarkers that operate as predictors for the metabolic modules. See ESF Statistics for further details on how regressions and machine learning techniques were performed.

#### Group level profiles

We employed PLS-DA, OPLS-DA, random forest, and SVM to investigate multivariate differentiation between MDD and control subjects. PLS-DA, OPLS-DA, and random forest were utilized to assess the importance of biomarkers in this discrimination. In cross-validation approaches, all feature selection, model training, hyperparameter optimization, and interpretability processes were performed exclusively within the training set, while the testing samples were reserved entirely for final evaluation. Feature selection was carried out using the top features in both PLS-DA and random forest, univariate analysis, and lasso regression as well (see ESF, Statistics). The initial data set of 736 metabolites was reduced to 16 (see **Table 1**), and based on those top features, we built 6 functional metabolite profiles (Maes et al., 2025) (Maes, Niu et al. 2025).

#### Phenome profiles

In order to delineate the most important biomarkers of the phenome profiles (OSOD, current SI, ROI, and physiosomatic symptoms), we performed PLS regression and multiple regression analyses (after performing lasso CATREG regression). We utilized PLS-regression to predict phenome scores (OSOD, present SI, physiosomatic symptoms) in the aggregated training and testing sets.

#### Depression modeling

Partial least squares (PLS) SEM analysis was utilized to investigate the causal relationships among the NIMETOX biomarkers, the metabolomics modules, and the clinical outcome. The latter was defined as a factor derived from OSOD, physiosomatic symptoms, ROI, and current SI, referred to as the “clinical phenome” (Niu, Zhang et al. 2025). Consequently, we constructed a latent vector representing metabolic disorders using metabolomics modules. Upon meeting specified model quality data criteria (see ESF, Statistics), we conduct a PLS-SEM path analysis using 5,000 bootstrap samples, deriving path coefficients (with associated p-values), and additionally calculating both specific and total indirect (mediated) and total effects.

#### Personalized profiles

PLS-regression was employed to obtain case-specific contribution scores that quantify the extent to which distinct biomarker profiles drive differentiation between MDD and control subjects, as well as the prediction of phenome scores. Scores for individual subjects on the PLS-derived latent components indicate each case’s location along the biomarker–phenome axes. The case-specific ratings (score-contribution scores) reflect the degree (relative predictive power) to which an individual’s biomarker profile drives MDD.

## Results

### Demographic, clinical and metabolomics data

Electronic Supplementary File (ESF) Table 2 shows the demographic and clinical data as previously described (Maes, Niu, et al., 2025). There were no significant differences in age, sex distribution, BMI, MetS prevalence, and education between MDD inpatients and controls. MDD patients showed increased OSOD, physiosomatic, suicidal ideation and ROI scores as compared with controls. ESF, Table 2 shows the mean values of these six functional modules in MDD inpatients and healthy controls. Lipotoxicity, PL remodeling, Mito ATP Redox, and FA storage/signaling were significantly higher in MDD versus controls, whereas ether lipids and Retinoid detox were significantly lower in MDD compared with controls.

**Table 2.**
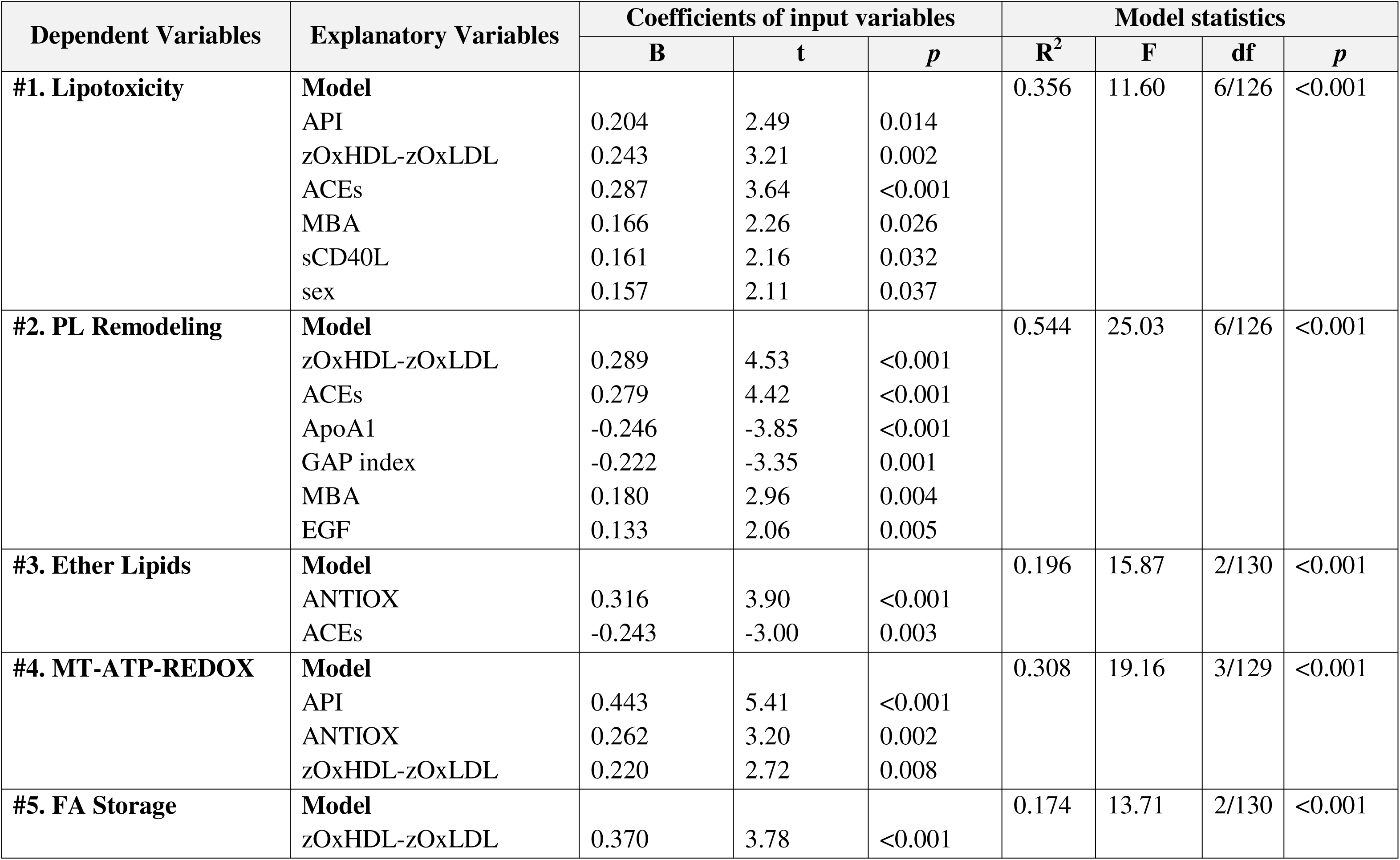
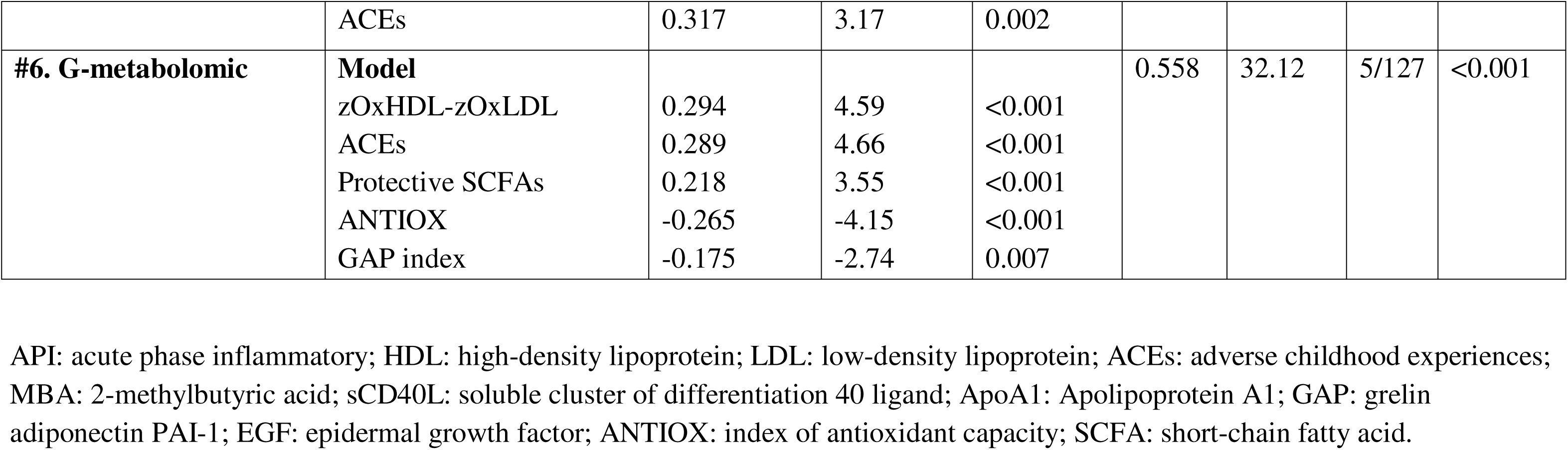
Results of multiple regression analysis with the overall severity of illness (OSOD), physiosomatic symptoms, current suicidal ideation (SI), and recurrence of illness (ROI) as dependent variables, and metabolites or functional module scores as explanatory variables.

| Dependent Variables | Explanatory Variables | Coefficients of input variables |  |  | Model statistics |  |  |  |
| --- | --- | --- | --- | --- | --- | --- | --- | --- |
|  |  | B | t | p | R<sup>2</sup> | F | df | p |
| #1. Lipotoxicity | **Model** |  |  |  | 0.356 | 11.60 | 6/126 | <0.001 |
|  | API | 0.204 | 2.49 | 0.014 |  |  |  |  |
|  | zOxHDL-zOxLDL | 0.243 | 3.21 | 0.002 |  |  |  |  |
|  | ACEs | 0.287 | 3.64 | <0.001 |  |  |  |  |
|  | MBA | 0.166 | 2.26 | 0.026 |  |  |  |  |
|  | sCD40L | 0.161 | 2.16 | 0.032 |  |  |  |  |
|  | sex | 0.157 | 2.11 | 0.037 |  |  |  |  |
| #2. PL Remodeling | **Model** |  |  |  | 0.544 | 25.03 | 6/126 | <0.001 |
|  | zOxHDL-zOxLDL | 0.289 | 4.53 | <0.001 |  |  |  |  |
|  | ACEs | 0.279 | 4.42 | <0.001 |  |  |  |  |
|  | ApoA1 | -0.246 | -3.85 | <0.001 |  |  |  |  |
|  | GAP index | -0.222 | -3.35 | 0.001 |  |  |  |  |
|  | MBA | 0.180 | 2.96 | 0.004 |  |  |  |  |
|  | EGF | 0.133 | 2.06 | 0.005 |  |  |  |  |
| #3. Ether Lipids | **Model** |  |  |  | 0.196 | 15.87 | 2/130 | <0.001 |
|  | ANTIOX | 0.316 | 3.90 | <0.001 |  |  |  |  |
|  | ACEs | -0.243 | -3.00 | 0.003 |  |  |  |  |
| #4. MT-ATP-REDOX | **Model** |  |  |  | 0.308 | 19.16 | 3/129 | <0.001 |
|  | API | 0.443 | 5.41 | <0.001 |  |  |  |  |
|  | ANTIOX | 0.262 | 3.20 | 0.002 |  |  |  |  |
|  | zOxHDL-zOxLDL | 0.220 | 2.72 | 0.008 |  |  |  |  |
| #5. FA Storage | **Model** |  |  |  | 0.174 | 13.71 | 2/130 | <0.001 |
|  | zOxHDL-zOxLDL | 0.370 | 3.78 | <0.001 |  |  |  |  |
|  | ACEs | 0.317 | 3.17 | 0.002 |  |  |  |  |
| **#6. G-metabolomic** | **Model** |  |  |  | 0.558 | 32.12 | 5/127 | <0.001 |
|  | zOxHDL-zOxLDL | 0.294 | 4.59 | <0.001 |  |  |  |  |
|  | ACEs | 0.289 | 4.66 | <0.001 |  |  |  |  |
|  | Protective SCFAs | 0.218 | 3.55 | <0.001 |  |  |  |  |
|  | ANTIOX | -0.265 | -4.15 | <0.001 |  |  |  |  |
|  | GAP index | -0.175 | -2.74 | 0.007 |  |  |  |  |

API: acute phase inflammatory; HDL: high-density lipoprotein; LDL: low-density lipoprotein; ACEs: adverse childhood experiences; MBA: 2-methylbutyric acid; sCD40L: soluble cluster of differentiation 40 ligand; ApoA1: Apolipoprotein A1; GAP: grelin adiponectin PAI-1; EGF: epidermal growth factor; ANTIOX: index of antioxidant capacity; SCFA: short-chain fatty acid.

### Construction of “G-metabolomics”

To construct a generalized latent vector (feature reduction), reflecting metabolic aberrations, we used PCA and extracted the first PC from the 16 top metabolomics that were delineated in the training sample (Maes, Niu et al. 2025). The first factor loaded highly on 10 of the 16 metabolites, and consequently, PCA was performed on those 10 variables. The KMO value was adequate (0.893) as was the Bartlett’s test of sphericity (χ^2^ = 1189.808, df = 45, *p* < 0.001). This first PC explained 57.26% of the variance in the data set. **Table 1** shows the loadings for these 10 metabolites, which cover 5 out of the 6 modules, all except Retinoid Detox. This first PC was labeled generalized (G)-metabolomics. ESF, Table 2 shows that the mean PC score was significantly higher in patients than controls.

### Relationships between metabolomic modules and other NIMETOX and metabolic features

Figure 1 shows a heatmap displaying the associations between the 7 metabolomic modules and selected key NIMETOX biomarkers. The G-metabolics, lipotoxicity, PL remodeling, ether lipids, and FA storage modules were significantly associated with most biomarkers. The Retinoid detox module, on the other hand, showed significant associations with two biomarkers only.

**Figure 1.**
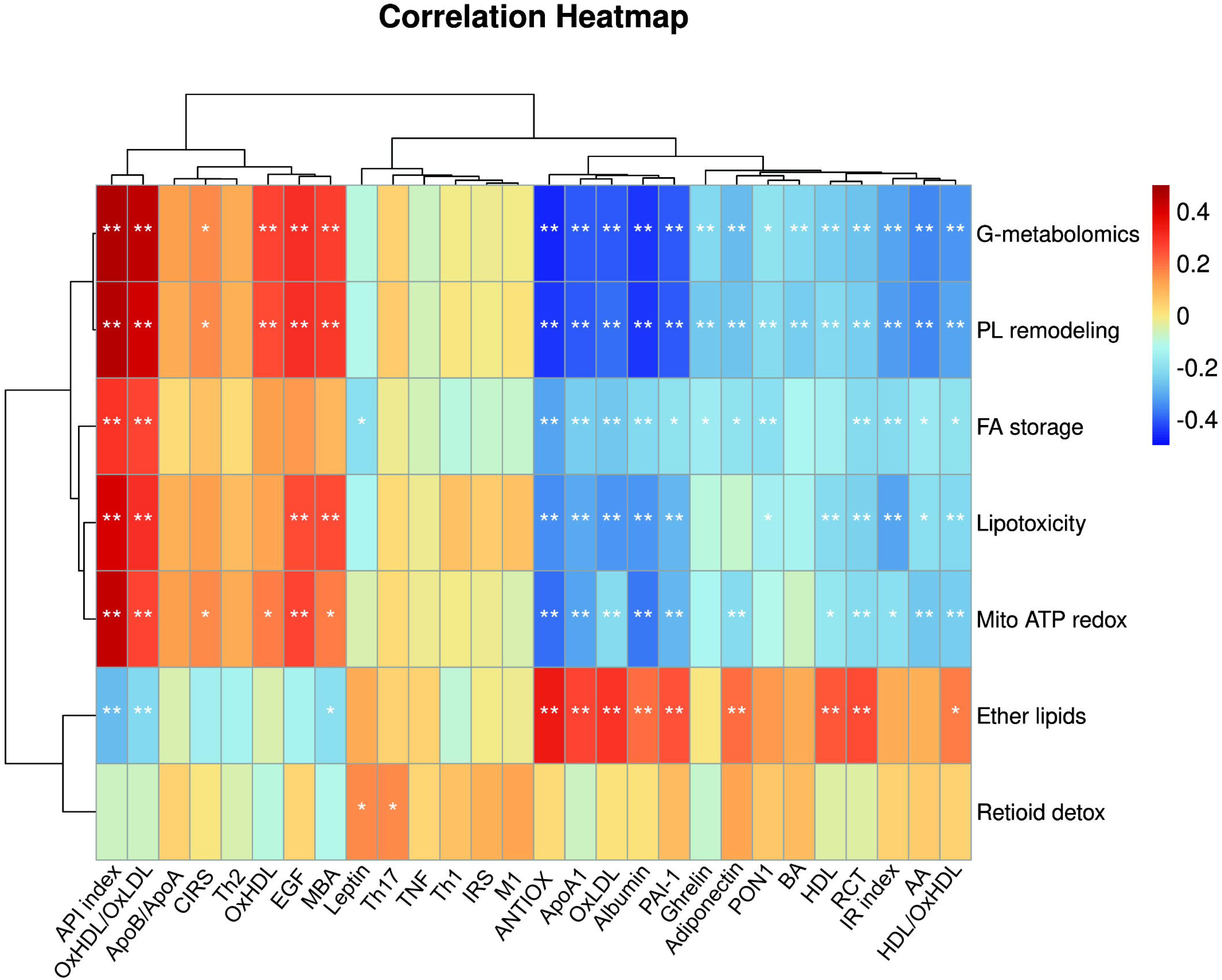
Heatmap displaying the associations between the 7 metabolomic modules and selected key NIMETOX biomarkers. See Table 1 and ESF, Table 2 for abbreviations and explanations about the biomarkers.

**Table 2** shows the results of multiple regression analysis with the 7 metabolic modules as dependent variables and the other biomarkers as explanatory variables. Regression #1 shows that 35.6% of the variance in lipotoxicity was explained by acute-phase inflammatory (API) index, zOxHDL-zOxLDL, ACEs, MBA, sCD40L, and male sex (all positively associated). Up to 54.4% of the variance in PL remodeling was explained by zOxHDL-zOxLDL, ACEs, MBA, EGF (all positively associated) and ApoA1 and the GAP index (both inversely associated). A smaller part of the variance in the ether lipids, ATP-Redox and FA storage modules was explained by ANTIOX, ACEs, zOxHDL-zOxLDL and/or API. We found that 55.8% of the variance in G-metabolomics was associated with zOxHDL-zOxLDL, ACEs, MBA (all positively associated) and ANTIOX, GAP index, and protective SCFAs (all inversely associated).

Table 2 of the ESF indicates that the prevalence of MetS was similar among individuals with MDD (17.7%) and controls (25.6%), and that there were no statistically significant differences in BMI and waist circumference between inpatients and controls. Figure 2 presents a heatmap illustrating correlations between the seven metabolic modules and BMI, MetS, as well as age and sex. We did not observe any significant associations among these variables, except for two trends, namely BMI and lipotoxicity (*p* = 0.018), and MetS and FA storage (*p* = 0.049). These significances were no longer observed following the FDR p-value correction. It should be emphasized that we consistently included MetS and BMI in the multiple regression analysis outlined below, and that neither BMI nor MetS achieved statistical significance. Our findings indicate that 92 patients with MDD were administered antidepressants, 68 were prescribed benzodiazepines, 44 received atypical antipsychotics, and 10 were provided with mood stabilizers. These medications did not yield any substantial impact on the 16 metabolites or the 7 domains, even in the absence of FDR p-value correction. Consequently, there is no evidence suggesting that the patient’s medication status affected the study outcomes.

**Figure 2.**
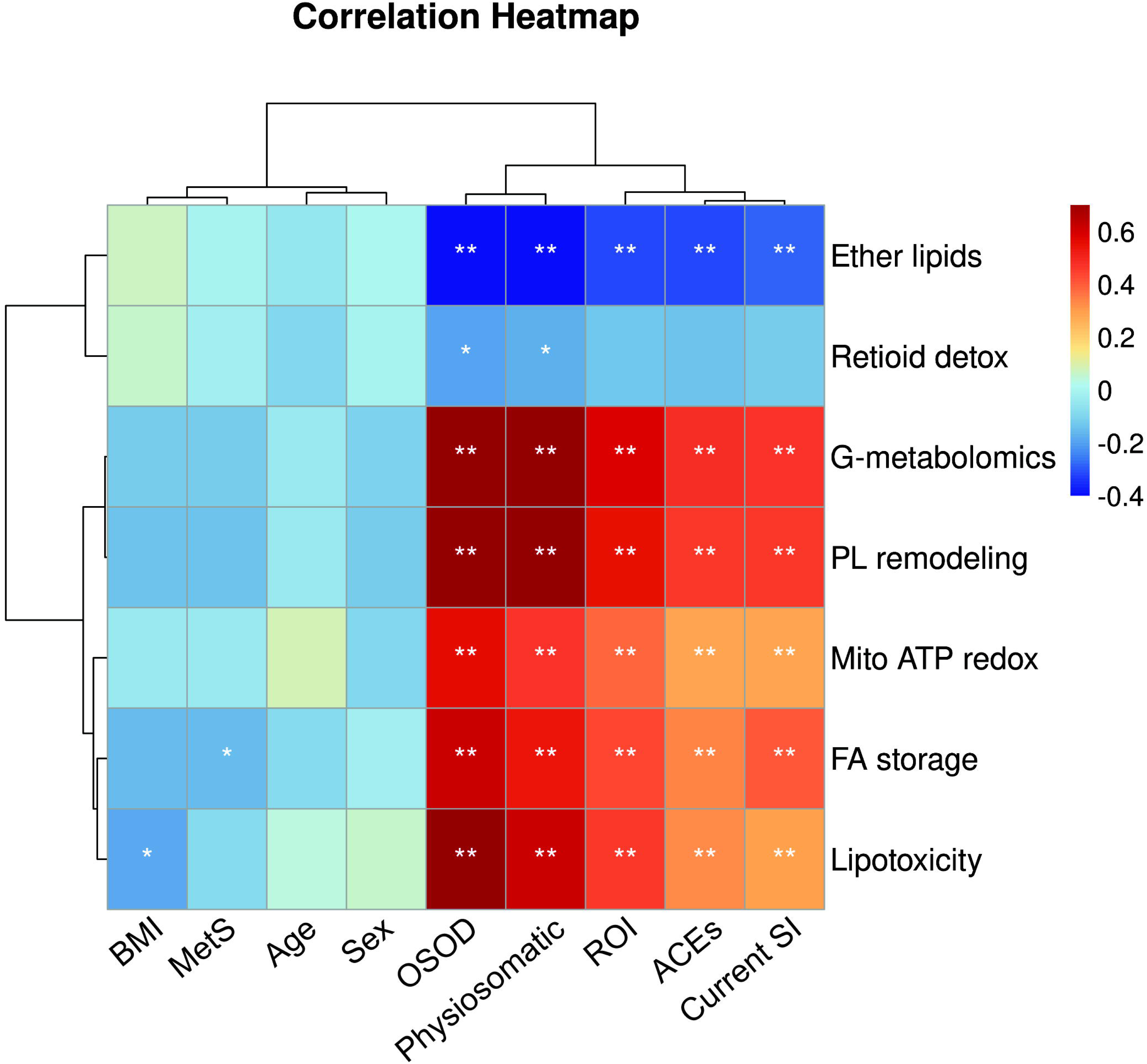
Heatmap showing the correlations between the seven metabolic modules and body mass index (BMI), metabolic syndrome (MetS), age, sex, and clinical data. See Table 1 for abbreviations and explanations about the metabolomics biomarkers. OSOD: overall severity of depression; Physiosomatic symptoms; ROI: recurrence of illness; ACEs: adverse childhood experiences; SI: suicidal ideation.

### Group level analysis

Partial least squares–discriminant analysis (PLS-DA), including all biomarkers (6 metabolic modules + all biomarkers), shows significant discrimination between MDD and controls. PLS-DA incorporating the six metabolomic modules together with the additional NIMETOX biomarkers, yielded a robust multivariate separation. The maximum number of NIPALS iterations was set to 50, with a convergence criterion of 0.001. The optimal number of components was determined by 7-fold cross-validation, which was also used to estimate model performance. Three latent components were retained, explaining a cumulative R²Y of 0.8726, with a cumulative Q² of 0.4862 and R²X of 0.2819 after two iterations, indicating good explanatory capacity and acceptable predictive performance. The eigenvalues of the three components were 8.439, 2.631, and 2.503, respectively. Figure 3 displays the VIP scores for all biomarkers, grouped into five functional categories: neuroimmune, metabolic, oxidative stress, gut microbiome, and basic clinical/biochemical variables. Sixteen variables showed VIP > 1, ranked by importance, led by lipotoxicity, PL remodeling, FA storage, Mito ATP redox stress, and antioxidant defenses, followed by acute phase/inflammation, ether lipids, ApoA1, ACEs, albumin, arachidonic acid, OxHDL/OxLDL indices, HDL/OxHDL ratio, PAI, EGF, and RCT.

**Figure 3.**
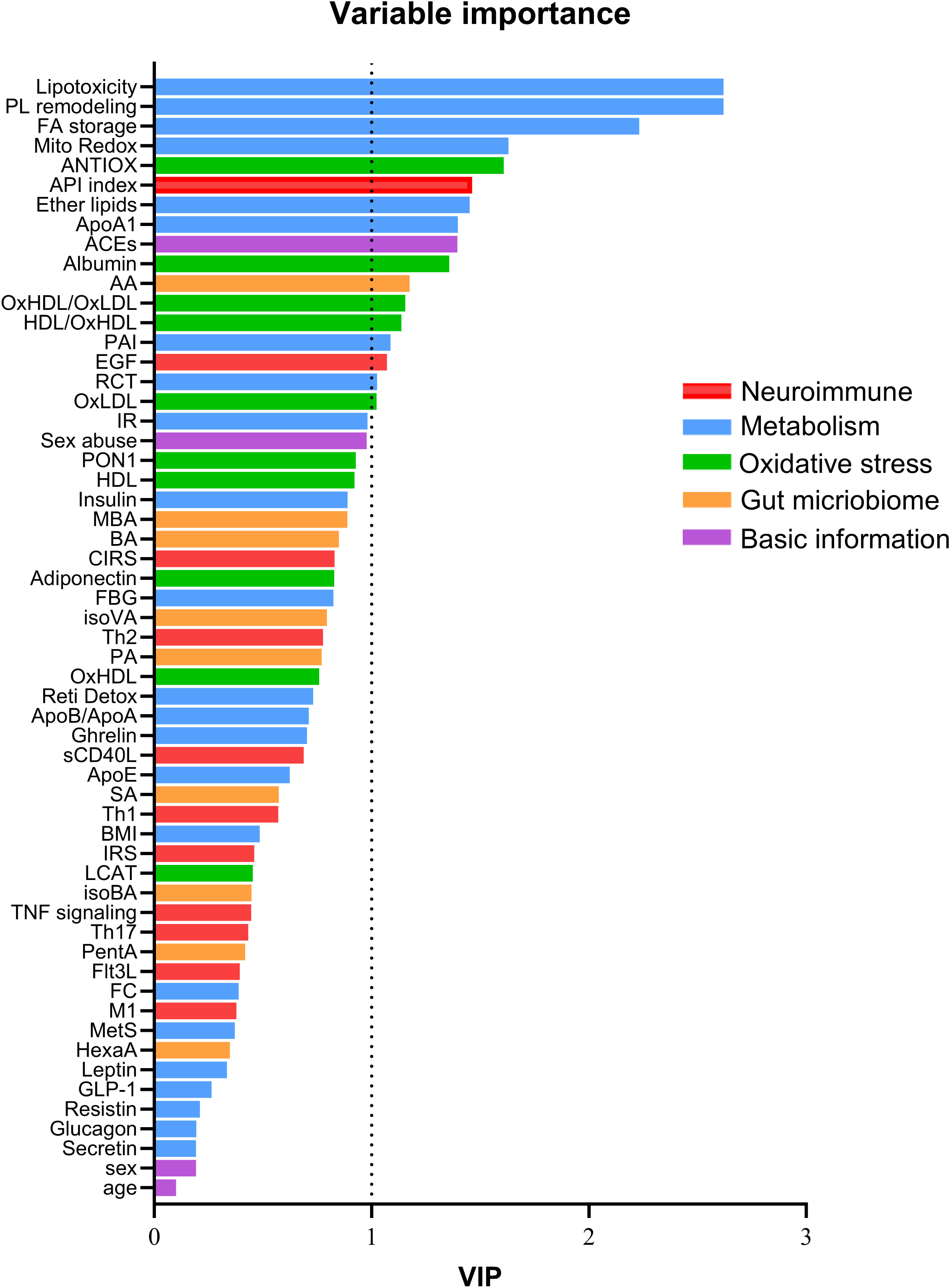
Results of partial least squares discriminant analysis (PLS-DA), which show the variable importance in projection (VIP) scores for all biomarkers discriminating inpatients with severe major depressive disorder from healthy controls. See Table 1 and ESF, Table 2 for abbreviations and explanations about the biomarkers. The biomarkers are grouped into five functional categories: neuroimmune, metabolic, oxidative stress, gut microbiome, and basic clinical variables, including adverse childhood experiences (ACEs), sexual abuse, sex, and age. BMI: body mass index.

Random forest analysis showed adequate classification performance. Using 70 trees, the model achieved 100% accuracy in the training sample, indicating complete separation between MDD patients and healthy controls. In the testing sample, random forest yielded a sensitivity of 97.9% and a specificity of 94.4%, demonstrating strong generalizability. Variable-importance analysis identified the top 10 predictors, ranked in descending order, as lipotoxicity, PL remodeling, FA storage, API index, retinoid detoxification, antioxidant defenses, RCT, glucose-dependent insulinotropic peptide (GIP), ACEs, and HDL-related measures.

SVM classification demonstrated strong discriminative performance. In the training sample, the model achieved 100% accuracy, indicating complete separation between MDD patients and healthy controls. In the testing sample, the classifier showed a sensitivity of 97.9% and a specificity of 83.3%, reflecting adequate detection of MDD with acceptable discrimination of controls. The model used classification type 1 with a radial basis function (RBF) kernel, where C = 4 provided a balanced trade-off between margin maximization and tolerance of classification errors, thereby reducing overfitting. The gamma parameter (γ = 0.017) defined a relatively smooth decision boundary, appropriate for high-dimensional metabolomic data. The presence of 58 support vectors, including 37 bounded vectors, indicates that classification relied on observations close to the decision boundary, consistent with a complex but stable separation. The decision was constant 0.375119, indicating a stable separating hyperplane with good generalization to unseen data.

Subsequently, biomarkers showing weak or noisy signals in the training sample were excluded. Feature reduction was guided by variable importance metrics, known collinearity between predictors, and stability across models. This step aimed to reduce redundancy, improve model robustness, and minimize overfitting. The refined feature set yielded a more parsimonious and accurate predictor panel with improved generalizability to the testing sample. In addition, to prevent blocks of highly correlated variables—particularly the six metabolomic modules—from disproportionately loading on the PLS-DA components and thereby obscuring other relevant predictors, we used only 3 metabolomic modules. This approach reduced dominance effects and improved the interpretability of the multivariate structure. The composite biomarkers were derived from the top-ranking variables identified in the training sample using ANOVA, PLS-DA, OPLS-DA, and random forest importance. This strategy ensured a more balanced representation of metabolic information while preserving discriminative power.

Figure 4A/B shows the results of PLS-DA and OPLS-DA, which revealed a clear separation between the two study groups. In PLS-DA, the first latent component (t) explained 36.12% of the variance relevant to group discrimination, while the second component (t) accounted for an additional 12.83%, together capturing nearly half of the discriminatory information in the dataset. OPLS-DA exhibited strong group differentiation, with the first predictive component accounting for 34.9% of the variance associated with class separation, while the second orthogonal component accounted for 14.7% of variation not connected to class membership. The OPLS-DA model demonstrated robust explanatory and predictive capabilities (R²Y = 66.9%, Q² = 64.8%), both statistically significant (p = 0.005), signifying a stable and non-overfitted discriminant model. Misclassifications in the 2D plots were limited to few cases. Figures 4C and **4D** show the importance of the 15 selected biomarkers for MDD versus controls using PLS-DA (4C) and Random Forest (4D), respectively. The latter showed an accuracy of 96.2% in the training sample, whilst in the testing sample, the sensitivity was 100% with a specificity of 88.9%. Both random forest and PLS-DA showed a similar profile with Lipotoxicity, PL modeling, ANTIOX, API, Retinoid Detox, and ACEs as the top features for MDD.

**Figure 4.**
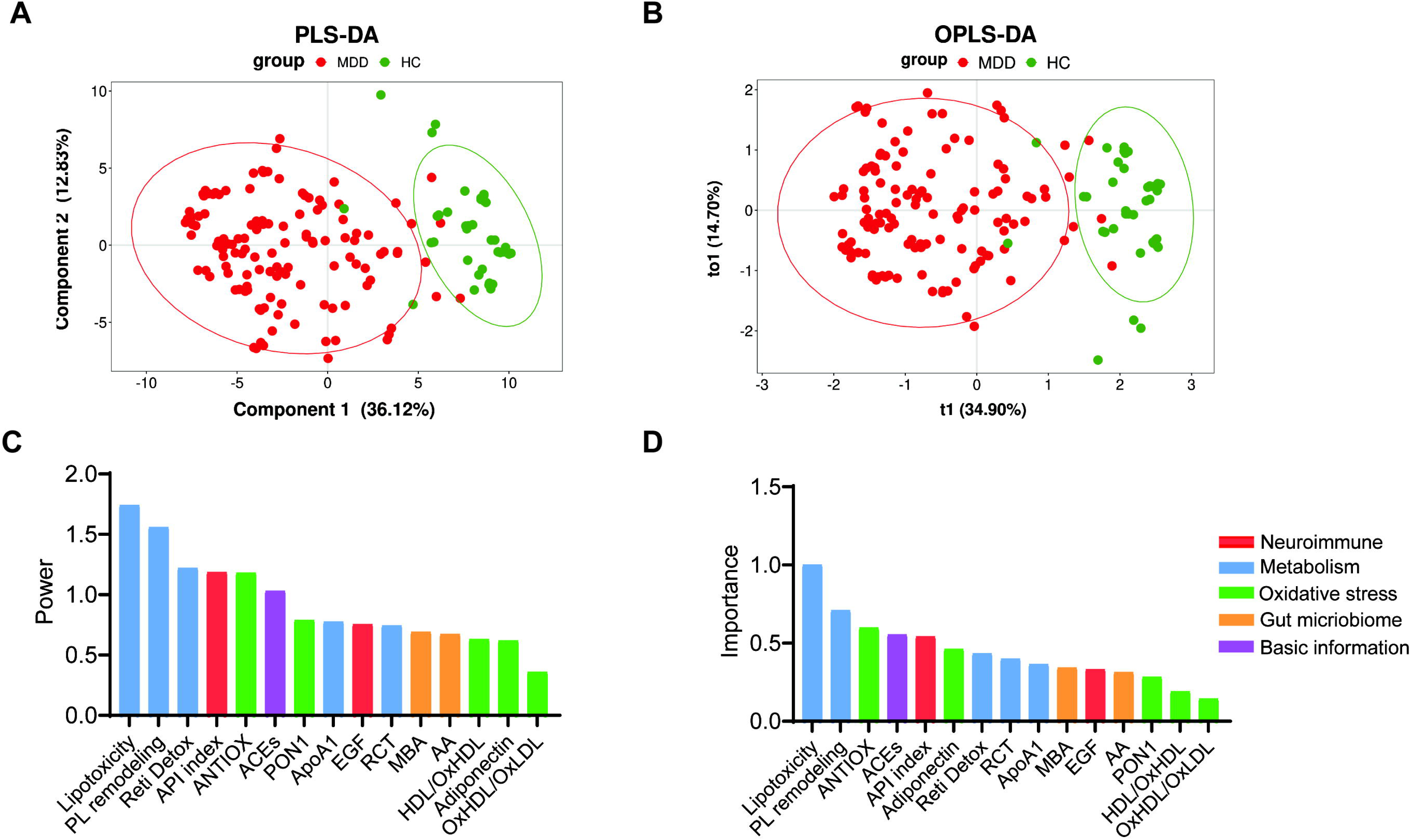
Results of partial least squares discriminant analysis (PLS-DA, Figure 4A) and orthogonal PLS-DA (OPLS-DA; Figure 4B). Both figures 4A and 4B show a clear demarcation between major depressive disorder (MDD) and healthy controls (HCs). In both analyses, the latent components account for nearly half of the discriminatory information in the dataset. Figure 4C shows the power (the variables’ contribution to the discriminant components obtained by PLS-DA using the 15 selected biomarkers. Figure 4D shows the results of Random Forest analysis (importance metrics) performed on the same 15 biomarkers. Both methods discriminated MDD inpatients from HCs with great accuracy. See Table 1 and ESF, Table 2 for abbreviations and explanations about the biomarkers.

### Phenome profiles

PLS-regression analyses were performed using the above 15 biomarkers as explanatory variables and the phenome data as dependent variables (Figure 5). For OSOD, three significant components were extracted, yielding an R²Y of 0.7699, R²X of 0.5367, and a cumulative Q² of 0.6601, indicating strong explanatory and predictive performance (Figure 5A). The most influential biomarkers (power > 1, indicating important predictors of the dependent variable), ranked in descending order, were PL remodeling, lipotoxicity, ANTIOX, ACEs, ApoA1, and API index. Moderate contributors (power between 0.8 and 1.0) were AA, HDL-OxHDL, and RCT. PLS regression of physiosomatic symptoms (Figure 5B) on the same biomarker predictors showed a comparable pattern, with the same five powerful and three moderate predictors and robust model performance (R²Y = 0.6399; R²X = 0.5366; Q² = 0.5145). Figure 5C shows the prediction of ROI (cumulative R²Y = 0.545; R²X = 0.4421; Q² = 0.4416). Figure 5D illustrates the PLS regression results for current suicidal ideation (one component was retrieved with R²Y = 0.3972; R²X = 0.4196; Q² = 0.2533), where the leading predictors were ACEs, PL remodeling, DAG lipotoxicity, ANTIOX, and ApoA1.

**Figure 5.**
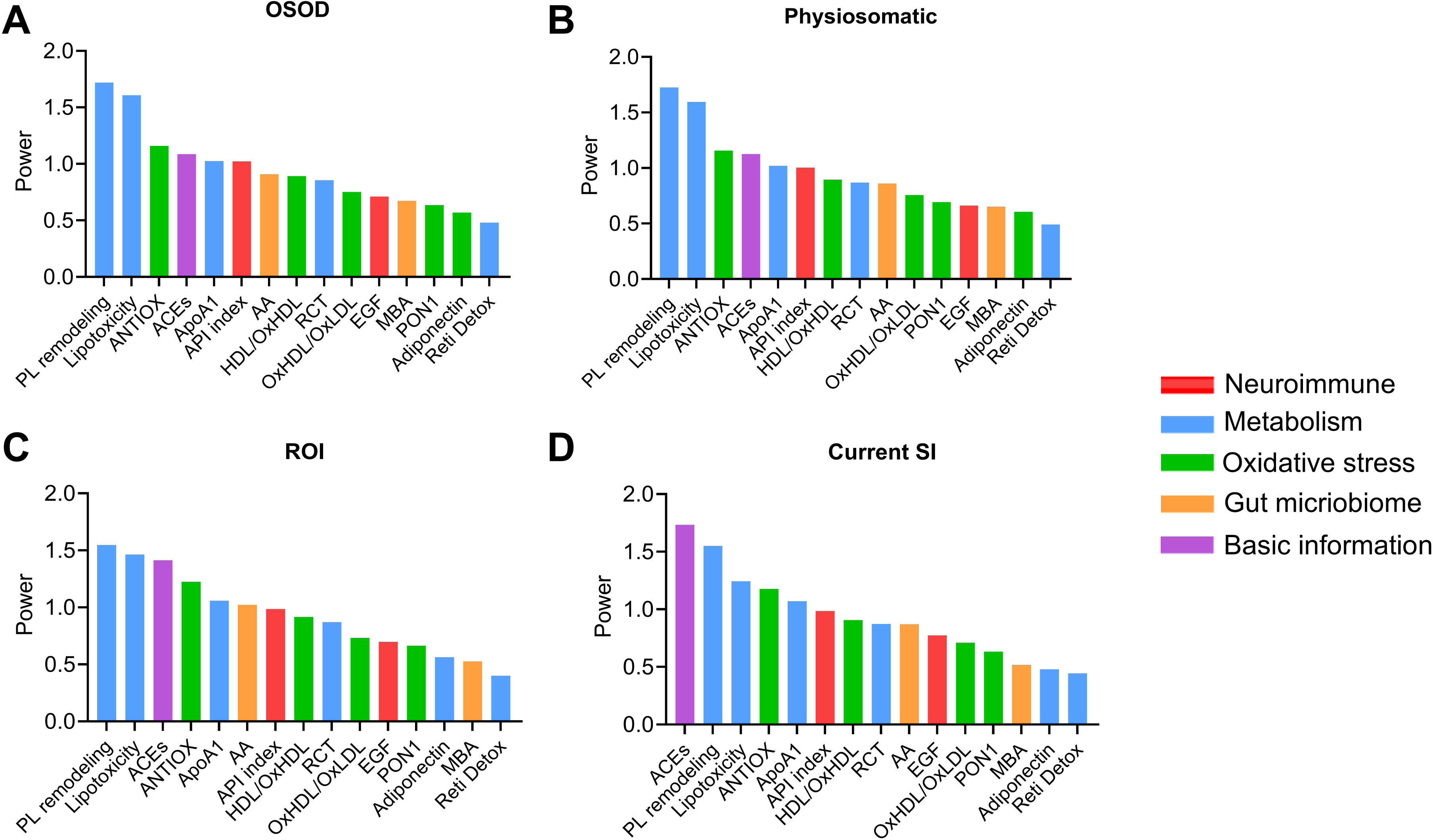
Results of partial least squares (PLS) regression analysis with the overall severity of depression (OSOD), physiosomatic symptom domain, recurrence of illness (ROI), and current suicidal ideation (SI) as dependent variables and 15 selected biomarkers as explanatory variables. See Table 1 and ESF, Table 2 for abbreviations and explanations about the biomarkers.

**Table 3** shows the results of multiple regression analysis of phenome features on biomarkers. Together, PL remodeling, FA storage, lipotoxicity, ACEs, Th17 (positively correlated) and ANTIOX (inversely) accounted for 76.3% of the variance in OSOD in the total study group. In patients with MDD, the variance in OSOD was largely attributable to PL remodeling, ACEs, Th2 (positively) and GIP (inversely), which together explained 24.3% of its variance. A multiple regression model including PL remodeling, Lipotoxicity, FA storage, sexual abuse (positively) and ANTIOX (inversely) explained 61.9% of the variability in physiosomatic symptoms in the total study group, In the restricted MDD sample, the combined effects of sexual abuse and ANTIOX yielded an explained variance of 10.0%. Regression on ACEs, FA storage, TNF signaling (positively), ApoA1 and BMI (inversely) produced a model explaining 40.6% of the variance in current SI. In the restricted MDD group, 30.1% of the variance in current SI was captured by a five-predictor model comprising ACEs, TNF signaling, FA storage (positively), GLP and ApoA1 (inversely). We found that 28.8% of the ROI variance was positively associated with increases in ACEs and Th17 and decreases in ANTIOX.

**Table 3.** Results of multiple regression analysis with the overall severity of illness (OSOD), physiosomatic symptoms, current suicidal ideation (SI), and recurrence of illness (ROI) as dependent variables, and metabolites or functional module scores as explanatory variables.

| Dependent Variables | Explanatory Variables | Coefficients of input variables |  |  | Model statistics |  |  |  |
| --- | --- | --- | --- | --- | --- | --- | --- | --- |
|  |  | B | t | p | R<sup>2</sup> | F | df | p |
| #1a. OSOD | **Model** |  |  |  | 0.763 | 72.28 | 5/135 | <0.001 |
|  | PL remodeling | 0.394 | 5.51 | <0.001 |  |  |  |  |
|  | FA storage/signaling | 0.197 | 3.80 | <0.001 |  |  |  |  |
|  | Lipotoxicity | 0.244 | 4.03 | <0.001 |  |  |  |  |
|  | ANTIOX | -0.129 | -2.68 | 0.008 |  |  |  |  |
|  | ACEs | 0.140 | 2.92 | 0.004 |  |  |  |  |
|  | Th17 | 0.092 | 2.15 | 0.033 |  |  |  |  |
| #1b. OSOD in MDD only | **Model** |  |  |  | 0.243 | 7.93 | 4/99 | <0.001 |
|  | PL Remodeling | 0.230 | 2.51 | 0.014 |  |  |  |  |
|  | ACEs | 0.277 | 3.03 | 0.003 |  |  |  |  |
|  | Th2 | 0.233 | 2.59 | 0.011 |  |  |  |  |
|  | GIP | -0.196 | -2.22 | 0.028 |  |  |  |  |
| #2a. Physiosomatic symptoms | **Model** |  |  |  | 0.619 | 37.62 | 5/116 | <0.001 |
|  | PL remodeling | 0.333 | 3.50 | <0.001 |  |  |  |  |
|  | Lipotoxicity | 0.235 | 2.80 | 0.006 |  |  |  |  |
|  | FA storage/signaling | 0.197 | 2.75 | 0.007 |  |  |  |  |
|  | ANTIOX | -0.152 | -2.33 | 0.022 |  |  |  |  |
|  | Sexual abuse | 0.128 | 2.15 | 0.033 |  |  |  |  |
| #2b. Physiosomatic symptoms in MDD | **Model** |  |  |  | 0.100 | 5.60 | 2/101 | 0.005 |
|  | Sexual abuse | 0.234 | 2.47 | 0.015 |  |  |  |  |
|  | ANTIOX | -0.213 | -2.26 | 0.026 |  |  |  |  |
| #3a. Current SI | **Model** |  |  |  | 0.406 | 27.71 | 5/143 | <0.001 |
|  | ACEs | 0.355 | 5.07 | <0.001 |  |  |  |  |
|  | FA storage/signaling | 0.241 | 3.42 | <0.001 |  |  |  |  |
|  | ApoA1 | -0.217 | -3.08 | 0.003 |  |  |  |  |
|  | TNF signaling | 0.160 | 2.45 | 0.015 |  |  |  |  |
|  | BMI | -0.140 | -2.07 | 0.040 |  |  |  |  |
| **#3b. Current SI in MDD only** | **Model** |  |  |  | 0.301 | 8.96 | 5/104 | <0.001 |
|  | ACEs | 0.344 | 4.03 | <0.001 |  |  |  |  |
|  | TNF signaling | 0.144 | 1.71 | 0.091 |  |  |  |  |
|  | FA storage | 0.196 | 2.34 | 0.021 |  |  |  |  |
|  | GLP | -0.197 | -2.33 | 0.022 |  |  |  |  |
|  | ApoA1 | -0.180 | -2.13 | 0.036 |  |  |  |  |
| **#4. ROI in MDD only** | **Model** |  |  |  | 0.288 | 14.29 | 3/106 | <0.001 |
|  | ACEs | 0.442 | 5.22 | <0.001 |  |  |  |  |
|  | ANTIOX | -0.238 | -2.86 | 0.005 |  |  |  |  |
|  | Th17 | 0.189 | 2.35 | 0.027 |  |  |  |  |

ME0063834;Tetranor-12(S)-HETE; ME0148101;Cysteylhistidine; ME0111867;1-phosphatidyl-1D-myo-inositol 3-phosphate. PL: phospholipids; FA: fatty acids; GIP: gastric inhibitory polypeptide; ApoA1: Apolipoprotein A1; TNF: tumor necrosis factor; BMI: body mass index; GLP: glucagon-like peptide; ACEs: adverse childhood experiences; ANTIOX: index of antioxidant capacity

### Depression modeling

Figure 6 shows the first PLS model. The phenome was conceptualized as a latent vector extracted from OSOD, ROI, current SI, and physiosomatic symptoms (all loadings > 0.788, composite reliability = 0.927, EV = 0.770). This phenome factor was the final outcome and three factors (metabolism, immune activation, and HDL-ANTIOX), AA, MBA, API, GAP index, ACEs, sexual abuse, and sex served as input variables. Adequate factors could be extracted from 5 metabolomics functional modules (all loadings > 0.678, composite reliability = 0.851, EV = 0.584, labeled “metabolism”), five HDL-ANTIOX biomarkers (all loadings > 0.780, composite reliability = 0.959, EV = 0.813, labeled HDL-ANTIOX), and 4 cytokine profiles (all loadings > 0.780, composite reliability = 0.959, EV = 0.813, labeled “immune activation”). The SRMR of the saturated (0.044) and estimated (0.064) models were adequate. Confirmatory Tetrad Analysis showed that the latent vectors were not mis-specified as reflective models. PLS predict showed that the Q2 predicted values were all greater than zero. Discriminant validity was obtained for all variables, although the metabolism and phenome factors showed an HTMT ratio of 0.84. We found that 62.4% of the variance in the phenome was explained by Metabolism, HDL-ANTIOX, ACEs, and acetate. Together, ACEs, sexual abuse (risk factors), GAP index, AA, and HDL-ANTIOX (protective factors) accounted for 76.3% of the variance in “metabolism.” ACEs and the API response (risk factors) accounted for 17.0% of the variance in HDL-ANTIOX. ESF, Table 3 shows the specific indirect effects, while the total indirect effects are shown in ESF, Table 4, and the total effects in ESF, Table 5. Immune activation, API, GAP index, acetate, and MBA, HDL-ANTIOX, and metabolism had significant effects on the phenome. ACEs and sexual abuse significantly impacted the phenome via various mediator pathways (e.g., metabolism, GAP-index, AA, and MBA for ACEs; metabolism for sexual abuse). Interestingly, female sex increased both ACEs and sexual abuse and via mediator pathways impacted the phenome (higher in females than males).

**Figure 6.**
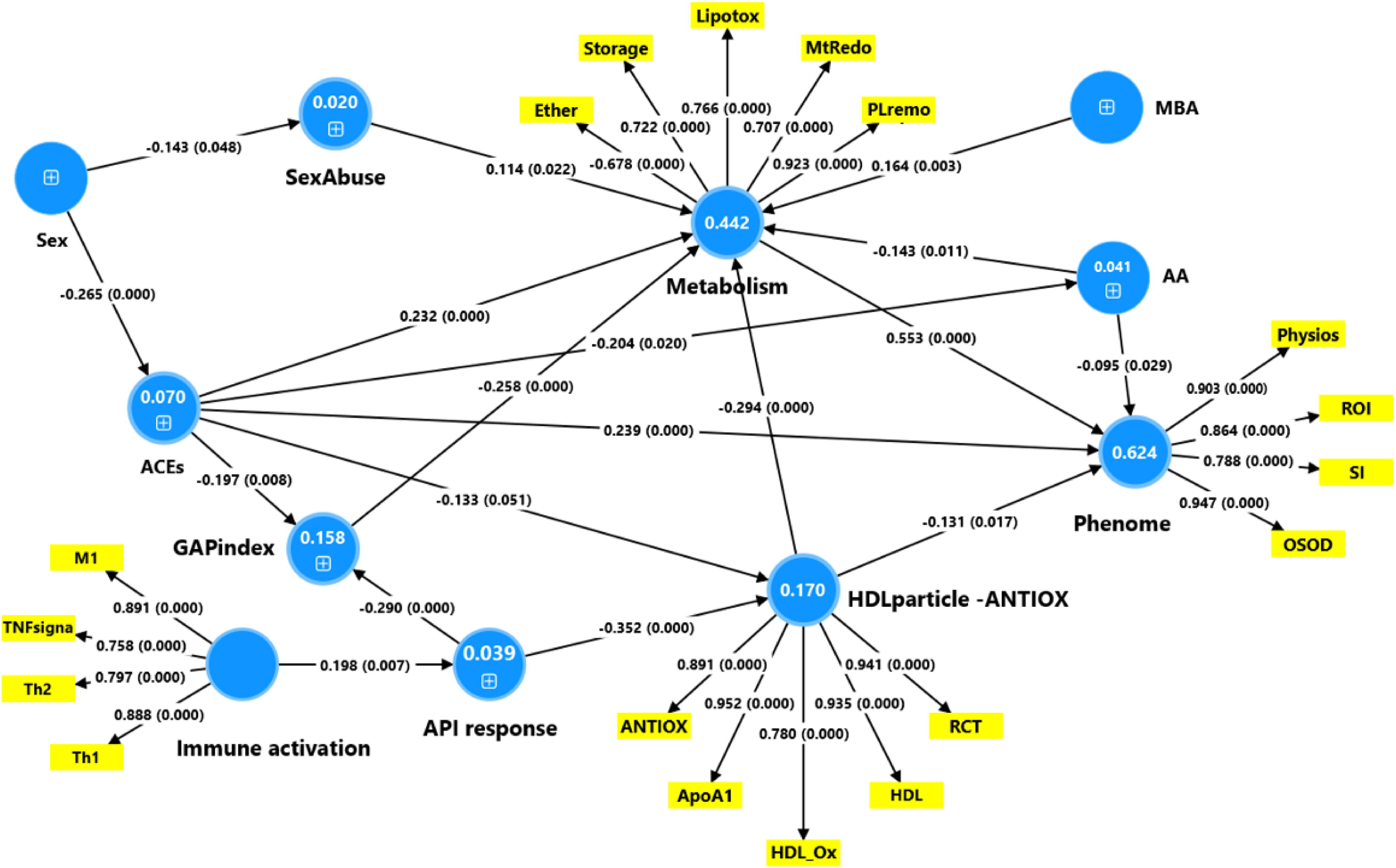
Results of partial least squares (PLS)-SEM analysis with the phenome latent vector as the final outcome variable and a metabolism latent vector, HDLparticle-ANTIOX latent vector, gut-derived acetate, and adverse childhood experiences (ACEs) as direct explanatory variables. The latter explains 62.4% of the variance in the phenome latent vector. ACEs, sexual abuse (sexAbuse), the HDL particle-ANTIOX latent vector, and the GAP (gherlin-adiponectin-PAI1) index explained 44.2% of the variance in the metabolism factor. The acute phase inflammatory (API) response is associated with the GAP index and HDL particle-ANTIOX factor and is predicted by immune activation. The phenome factor is conceptualized as a factor extracted from the overall severity of depression (OSOD), recurrence of illness (ROI), suicidal ideation (SI) and physiosomatic symptoms. The metabolism factor is extracted from 5 metabolomics modules, namely lipotoxicity (Lipotox), fatty acid storage (storage), phospholipid remodeling (PLremo), ether lipids (ether), and mitochondrial – Redox aberrations (MtRedo). The HDL particle-ANTIOX factor is extracted from total antioxidant defenses (ANTIOX), Apolipoprotein A1 (ApoA1), high-density lipoprotein cholesterol (HDL), oxidized HDL (HDL_ox), and reverse cholesterol transport (RCT) index. Immune activation is conceptualized as a factor extracted from M1 macrophages, tumor necrosis factor (TNF) signaling, T helper-1 (Th1) and Th2 immune phenotypes. See Table 1 and ESF, Table 2 for abbreviations and explanations about the biomarkers.

Figure 7 shows a second PLS model in which we successfully combined the three most important metabolomic modules with the phenome features. This new factor showed adequate metrics (all loadings > 0.685, composite reliability = 0.924, EV = 0.652, labeled “metabolic phenome”). The immune activation and HDL-ANTIOX factors were the same as in PLS model#1. In this model, we entered (instead of acetate) a reflective factor indicating “protective SCFAs” based on acetate, BA, and PA (all loadings > 0.879, composite reliability = 0.979, EV = 0.823). The SRMR of the saturated (0.041) and estimated (0.062) models were adequate. We found that 50.4% of the variance in the “metabolic phenome” was explained by lower levels of protective SCFAs, GAP index, and HDL-ANTIOX (protective factors) and increased levels of MBA, ACEs, and sexual abuse (risk factors). The effects of immune activation on the “metabolic-phenome” were mediated via the API response and HDL-ANTIOX. The effects of ACEs on the “metabolic-phenome” were mediated via HDL-ANTIOX, GAP index, API response, and protective SCFAs. ESF, Table 6 shows the total effects obtained in this PLS model #2 in descending order of importance: ACEs, HDL-ANTIOX, API index, protective SCFAs, sex, GAP index, MBA, cytokine networks, and sexual abuse.

**Figure 7.**
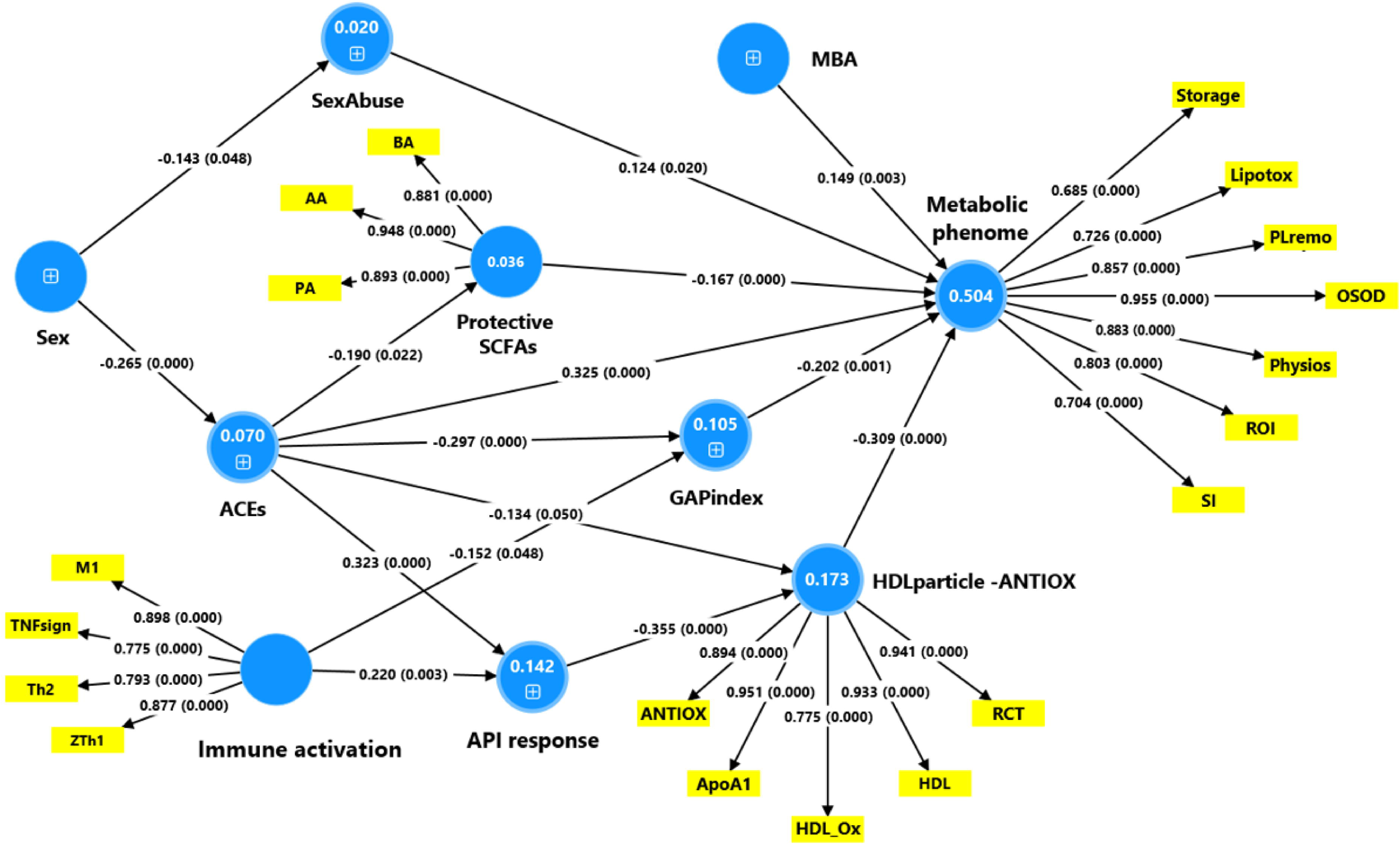
Results of a second partial least squares (PLS)-SEM analysis, which combined overall severity of depression (OSOD), physiosomatic symptoms, recurrence of illness (ROI) and suicidal ideation (SI) together with lipotoxicity (Lipotox), fatty acid storage (storage) and phospholipid remodeling (PLremo) in one factor. This factor is labeled the “metabolic phenome.” The latter is predicted by the HDL particle-ANTIOX factor, protective short-chain fatty acid (SCFA) factor, 2-methybutyrate (MBA), GAP (gherlin-adiponectin-PAI1) index, adverse childhood experiences (ACEs), and sexual abuse (SexAbuse). The HDL particle-ANTIOX factor is conceptualized as a factor extracted from total antioxidant defenses (ANTIOX), Apolipoprotein A1 (ApoA1), high-density lipoprotein cholesterol (HDL), oxidized HDL (HDL_ox), and the reverse cholesterol transport (RCT) index. The protective SCFA factor is conceptualized as the first factor extracted from butyrate (BA), acetate (AA), and propionate (PA). Immune activation is conceptualized as a factor extracted from M1 macrophages, tumor necrosis factor (TNF) signaling, Thelper-1 (Th1) and Th2 immune phenotypes. See Table 1 and ESF, Table 2 for abbreviations and explanations about the biomarkers.

### Personalized approach via PLS-regression

The PLS-DA analysis reported above, including 15 selected biomarkers, was used to calculate score-contribution profiles for each participant to demonstrate the relative predictive power of the model for MDD. Figure 8 illustrates sample profiles from four MDD inpatients.

**Figure 8.**
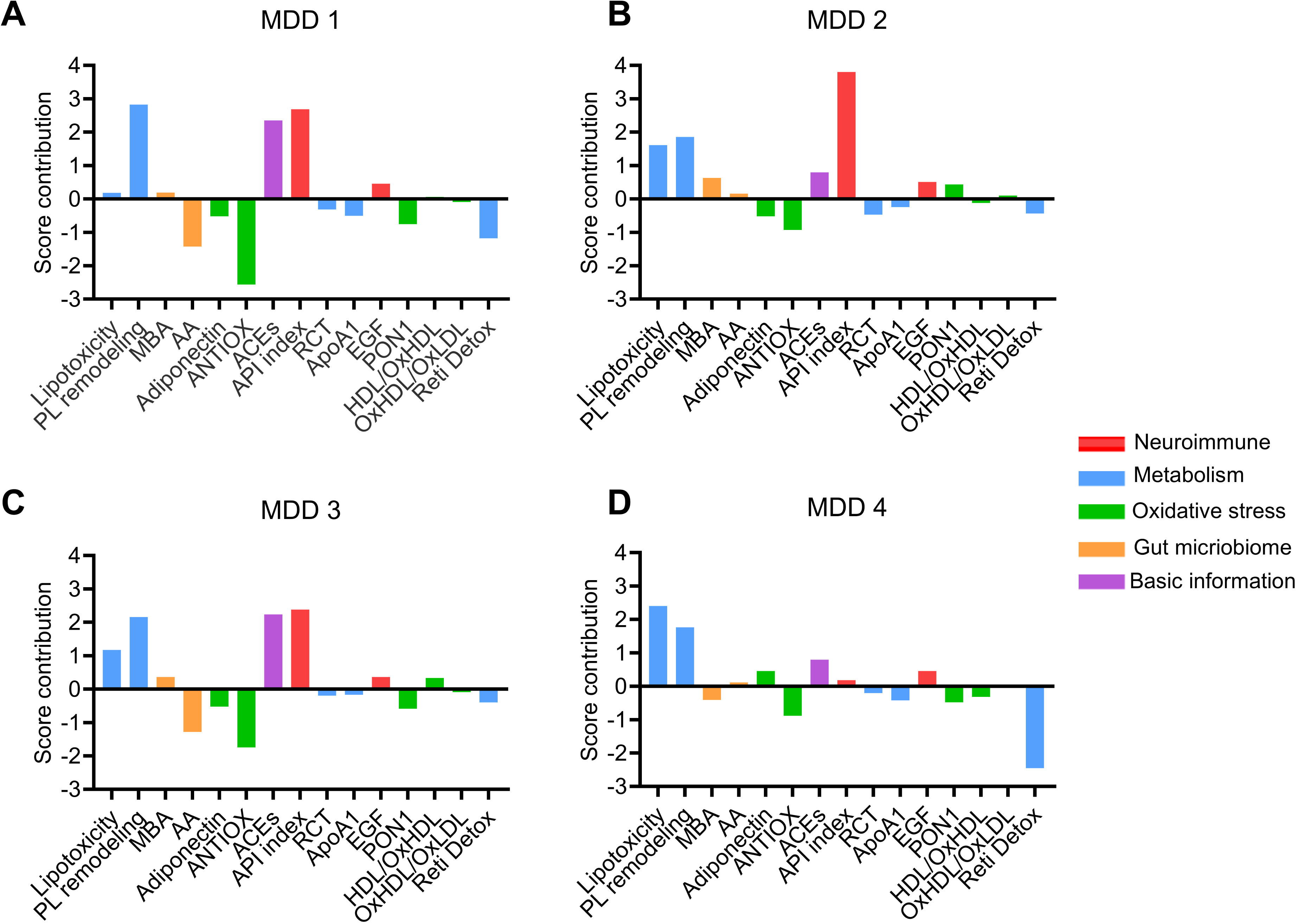
Score contribution profiles computed using partial least squares regression analysis (see Figure 5). This figure shows the personalized profiles based on relative power predicting severe major depressive disorder (MDD) in 4 different inpatients.

## Discussion

### The NIMETOX profiles underlying MDD

At the group level, patients with MDD were distinctly separated from healthy controls through an integrated NIMETOX profile that included metabolic alterations, immune activation, disturbances in SCFAs, changes in energy homeostasis metabolic hormones and adipokines, diminished antioxidant defenses, impaired RCT, decreased ApoA1 and PON1 activity, and increased HDL oxidation. This multimodal signature emphasizes that MDD is not characterized by a solitary biological anomaly but rather by a coordinated dysregulation across multiple neuroimmune, metabolic, endocrine, and other systems (Maes, Almulla et al. 2025, Maes, Jirakran et al. 2025).

Previous studies have demonstrated many of these abnormalities in isolation. For instance, it has been demonstrated that MDD is associated with neuro-immune activation, an acute phase response, diminished lipid-associated antioxidant defenses, and decreased levels of protective SCFAs and adiponectin (Maes, Vandewoude et al. 1991, Maes 1993, Maes, Smith et al. 1997, Maes, Galecki et al. 2011, Hu, Dong et al. 2015, Almulla, Thipakorn et al. 2023, Almulla, Niu et al. 2025, Liu, Geng et al. 2025, Maes, Almulla et al. 2025, Li, Murtaza et al. 2026). The current findings expand upon this body of literature by illustrating that these pathways converge into a consistent biological pattern that distinctly differentiates MDD from control groups.

Importantly, among all predictors, the metabolomic variables proved to be the most significant, especially those associated with lipotoxicity, PL remodeling, and FA storage and redistribution. The second most significant contributors to the discrimination of MDD were elements of the API response, combined with diminished antioxidant capacity, including decreased ApoA1 levels and PON1 activity. Furthermore, the intersections between the two metabolic pathways and redox processes are significantly implicated, as evidenced by decreased levels of ether lipids and retinoid detoxification. Together, these findings suggest that although immune and redox disturbances continue to be fundamental, metabolic reprogramming—particularly in the domains of lipid processing and membrane biology—serves a pivotal role in characterizing the MDD phenotype across populations. Indeed, the metabolomic profile of MDD corresponds to a consistent NIMETOX-driven biochemical phenotype. The pattern suggests the convergence of stress-induced lipolysis, arachidonic and adrenic membrane remodeling, mitochondrial redox overload, and depletion of antioxidant buffering capacity, all of which are integral to the NIMETOX model of MDD (Maes, Almulla et al. 2025) (Maes et al., 2025a).

### Phenotypic profiles and personalized fingerprints

Across phenotypic dimensions of MDD, including OSOD, physiosomatic symptom burden, ROI, and current suicidal ideation, outcomes were robustly predicted by pathways related to NIMETOX. Multivariate models consistently showed that the NIMETOX framework offers substantial explanatory capacity for clinical manifestations, correlating NIMETOX irregularities with symptom severity and disease progression.

Among all biological predictors, metabolomic variables were identified as the most significant determinants of these phenotypic characteristics. In particular, metabolomic domains associated with lipotoxicity, PL remodeling, FA storage/redistribution, and mitochondrial redox stress demonstrated the most robust and consistent correlations with OSOD, physiosomatic symptoms, suicidal ideation, and ROI. The immune-inflammatory components of NIMETOX, including the API response and reduced antioxidant levels, contributed substantially; however, their predictive power was subordinate to that of the metabolomic signatures.

Furthermore, our PLS-SEM analysis demonstrated that these three metabolic modules, OSOD, physiosomatic symptoms, SI, and ROI, represent manifestations of a common latent construct, which can be conceptualized as the “metabolic-phenome.” These findings indicate that metabolic reprogramming is not merely associated with MDD diagnosis but serves as a fundamental factor influencing symptom severity, suicidal behaviors, and the recurrence of illness. Together, these findings demonstrate that NIMETOX-informed biological insights can predict not only the occurrence of MDD but also its phenotypic heterogeneity, with metabolomics offering the most comprehensive explanatory framework across clinical parameters.

Significant heterogeneity was observed among the inpatients with MDD: some displayed considerable positive influences from PL remodeling, ACEs, and reduced antioxidant defenses in relation to the MDD diagnosis. Others exhibited an elevated API index or decreased Retinoid Detox levels accompanied by increased lipotoxicity. Thus, although a shared metabolomic signature of MDD is observed at the group level, each MDD patient displays a distinct biomarker profile, reflecting considerable interindividual metabolic heterogeneity in disease presentation. Since these NIMETOX biomarkers effectively distinguish MDD from controls, such personalized NIMETOX profiles serve as meaningful individualized identifiers that differentiate among MDD patients and between patients and controls (Maes and Stoyanov 2022, Maes, Moraes et al. 2023).

### Upstream NIMETOX pathways as causes of metabolic downstream findings

As demonstrated by PLS-SEM frameworks, NIMETOX may constitute the downstream, integrative manifestation of multiple upstream perturbations, rather than an isolated causal pathway. Immune activation, the API response, altered lipid transport (including decreased ApoA1, HDL, PON1, and RCT), diminished antioxidant defenses, metabolic hormone fluctuations, and SCFA imbalances may function upstream, converging to induce persistent nitro-oxidative lipid toxicity, membrane remodeling, and mitochondrial dysfunction. These convergent effects manifest as the NIMETOX phenotype, which subsequently influences metabolomic reprogramming and clinical presentation.

The primary contributors to the metabolic aberrations are diminished antioxidant defenses, primarily characterized by low HDL, ApoA1, PON1, impaired reverse cholesterol transport, and elevated OxHDL. This phenomenon may be attributed to the understanding that these antioxidant compounds inhibit the effective removal of oxidized phospholipids and saturated diacylglycerols, thereby maintaining a proinflammatory lipid environment (Morris, Puri et al. 2021, Almulla, Thipakorn et al. 2023). Low HDL levels may hinder the efflux of excess fatty acids, leading to increased intracellular fatty acid accumulation and facilitating DAG synthesis (Samuel and Shulman 2012). Simultaneously, reduced HDL levels diminish circulating ether lipids and plasmalogens, which are key endogenous antioxidants, thereby accounting for the decline in 1-O-hexadecyl-lysoPC, a marker of decreased ether-lipid antioxidant buffering (Stafforini 2009). Decreased PON1 activity results in elevated systemic ROS and lipid peroxidation, which in turn leads to aberrations in the Mito-ATP-Redox module (Gianazza, Brioschi et al. 2021).

Inadequate HDL-mediated removal of oxidized lipids perpetuates mitochondrial dysfunction (Maes, Galecki et al. 2011, Morris, Puri et al. 2021). Consequently, reductions in HDL/PON1 and the oxidation of HDL might lead to the accumulation of oxidized PL species, thereby contributing to PL remodeling and lipotoxicity (Durrington, Mackness et al. 2001, Gao and Podrez 2018, Maes, Niu et al. 2025). OxHDL itself exhibits proinflammatory properties by activating Toll-like receptor (TLR) 2/4 and NF-κB pathways in endothelial cells (Navab, Ananthramaiah et al. 2004), thereby promoting PLA_2_-LOX signaling and facilitating the production of eoxin C4 (Lin, Wang et al. 2025). Thus, the HDL/oxHDL–PON1 axis functions as a central regulator, enhancing oxidative membrane remodeling, DAG/PKC-induced lipotoxicity, leukotriene production, and mitochondrial redox stress, thereby comprehensively accounting for the metabolomic phenotype.

Reduced RCT impairs HDL-mediated clearance of oxidized phospholipids and excess membrane cholesterol, thereby facilitating PLA activation and the release of arachidonic and adrenic acids, which amplifies LOX-derived eicosanoid signaling (Tall and Yvan-Charvet 2015). Impaired RCT additionally disrupts endosomal phosphatidylinositol cycling and redirects fatty acids from inert storage toward bioactive phospholipid and DAG pools, thereby amplifying PKC-mediated inflammatory signaling. The accumulation of oxidized lipids further damages mitochondrial membranes, thereby elevating oxidative stress and promoting ATP degradation (Durrington, Mackness et al. 2001). These processes collectively contribute to the development of the metabolomic modules identified in MDD within the NIMETOX framework

ROS excess, resulting from mitochondrial abnormalities and diminished antioxidant capacity, accelerates the oxidation of PL species, leading to the formation of oxidized PLs and facilitating microparticle formation and blood-brain barrier (BBB) dysfunction (Gianazza, Brioschi et al. 2021). ROS-mediated proteolysis and amino acid redox cycling might account for the elevated levels of cysteinyldihistidine and the mitochondrial dysfunction marker hydroxyphenyllactic acid (Beloborodova, Pautova et al. 2019, Kehm, Baldensperger et al. 2021). Oxidative stress might also promote the turnover of retinoids, resulting in an increased formation of retinyl-acetate glucuronide (Penniston and Tanumihardjo 2006).

In fact, PS(22:5/22:4) may represent the most robust individual predictor of MDD, as it consolidates all main NIMETOX pathways—oxidative stress, cytokine activation, mitochondrial dysfunction, membrane remodeling, and HDL/PON1 impairment—into a highly sensitive structural indicator. This phosphatidylserine species comprises docosapentaenoic acid (22:5) and adrenic/arachidonic acid (22:4), two polyunsaturated fatty acids (PUFAs) highly susceptible to peroxidation, rendering it an early and preferential target of ROS/RNS-induced damage (Liang, Minikes et al. 2022, O’Donnell 2022). Its accumulation indicates PLA_2_–LOX activation, initiated by IL-1β, IL-6, and TNF-α, processes that are persistently elevated in MDD (Leslie 2015). PSs are predominantly localized in mitochondrial and endothelial membranes, and oxidative stress induces PS externalization on the outer leaflet, resulting in microparticle formation that indicates microvascular injury and BBB dysfunction (Hugel, Martínez et al. 2005, Bevers and Williamson 2016, Edrissi, Schock et al. 2016).

Our PLS-SEM model indicates that the API response is the second most significant contributor to the metabolomics module. Reduced levels of albumin and transferrin diminish plasma antioxidant buffering capacity, thereby elevating the availability of peroxidized lipids (Maes, Galecki et al. 2011). Albumin serves as the primary transporter and buffer for non-esterified fatty acids, oxidized lipids, and reactive aldehydes, and possesses significant thiol-dependent antioxidant properties (Roche, Rondeau et al. 2008, Mishra and Heath 2021). When albumin levels are decreased and undergo oxidative modification, saturated and polyunsaturated fatty acid species are less effectively buffered and are more readily diverted into PLs and DAGs (van der Vusse 2009, Blache, Bourdon et al. 2015).

Low transferrin levels elevate the pool of non-transferrin-bound iron, thereby promoting Fenton chemistry and the generation of hydroxyl radicals (Brissot, Ropert et al. 2012, Duca, Di Pierro et al. 2025). These reactive species oxidize PUFA-rich PLs and ether lipids, leading to the depletion of ether lipids and an increase in mitochondrial, metabolic and oxidative stress biomarkers such as HPLA (Beloborodova, Pautova et al. 2019).

Monomeric C-reactive protein (mCRP), in comparison to native pentameric CRP, exhibits enhanced endothelial and platelet-activating capabilities, facilitating PS exposure, complement activation, and microparticle dispersal (Almulla, Niu et al. 2025). In the context of decreased albumin and transferrin levels, mCRP interacts with membranes already enriched in arachidonic acid/adrenic PI/PS/PC and saturated DAGs, promoting additional externalization of PS, microthrombosis, and localized ROS production (Molins, Peña et al. 2008, Ji, Ma et al. 2009, Thiele, Habersberger et al. 2014). Thus, the combination of low albumin, low transferrin, and elevated mCRP eliminates critical antioxidant and metal-binding buffers, thereby exerting a pro-thrombo-inflammatory influence on the vasculature. This consolidates the observed pattern of DAG/PKC lipotoxicity, PL remodeling, leukotriene overproduction, mitochondrial redox stress, and retinoid detoxification activation in MDD.

Our findings indicate that the API index exerted a significantly greater influence on the MDD phenotype compared to various immune profiles (e.g., M1, Th1, TNF signaling). Nonetheless, the influence of these immune characteristics on the phenome was mediated by the API index. It is essential to recognize that EGF was a significant predictor of MDD and correlated with overall metabolic alterations. Activation of the EGF receptor (EGFR) directly stimulates PLA activity, increases phosphatidylinositol (PI) cycle turnover, and promotes DAG–PKC signaling, thereby strengthening phospholipid remodeling and lipotoxic membrane stress (Margolis, Holub et al. 1988). Meanwhile, EGFR signaling enhances mitochondrial metabolic requirements, accelerates oxidative phosphorylation, and increases ROS generation, potentially surpassing antioxidant defenses and intensifying mitochondrial redox imbalance (Bae, Kang et al. 1997, Orofiamma, Vural et al. 2022). Together, these effects establish a mechanistic connection between increased EGF signaling and the enhancement of the NIMETOX-associated metabolic modules.

Lower butyrate, acetate and propionate, coupled with elevated 2-MBA levels, suggest a transition from saccharolytic to proteolytic fermentation, which diminishes the anti-inflammatory, barrier-protective, and mitochondrial-supportive signaling typically mediated by BA (Morrison and Preston 2016). As a consequence, diminished BA levels may eliminate the tonic suppression of PLA /LOX and NF-κB, thereby enhancing LOX eicosanoid signaling and DAG–PKC signaling (Lee, Kim et al. 2017, Zhou, Ji et al. 2021). Simultaneously, elevated 2-MBA indicates proteolytic dysbiosis that facilitates oxidative stress and increased epithelial permeability, thereby indirectly exacerbating mitochondrial redox imbalance and antioxidant depletion across the six modules (Gilbert, Ijssennagger et al. 2018, Diether and Willing 2019). The above SCFA profile also suggests gut dysbiosis and increased intestinal permeability, which may lead to increased levels of lipopolysaccharide (LPS) and microbial metabolites (Maes, Kubera et al. 2008, Rudzki and Maes 2021). These processes may activate TLR4 and nucleotide-binding domain leucine-rich repeat and pyrin domain-containing receptor 3 (NLRP3) pathways (Maes, Almulla et al. 2025), thereby elevating IL-1β, IL-6, and TNF-α, which in turn stimulate the activity of PLA_2_ and LOX (Triggiani, Granata et al. 2005).

Finally, reduced adiponectin levels can further exacerbate metabolomic aberrations by diminishing crucial anti-inflammatory and lipid-buffering signals (Ajuwon and Spurlock 2005, Ouchi, Parker et al. 2011). Adiponectin typically inhibits PLA –arachidonic acid release, LOX/COX eicosanoid synthesis, and DAG–PKC signaling, while promoting mitochondrial efficacy; consequently, decreased adiponectin levels might contribute to lipotoxicity, phospholipid remodeling, and mitochondrial redox stress (Ajuwon and Spurlock 2005, Civitarese, Ukropcova et al. 2006, Ouchi, Parker et al. 2011, Li, Zhang et al. 2020). Adiponectin maintains intestinal barrier integrity and mitigates metabolic endotoxemia, while its decrease correlates with heightened epithelial permeability and inflammatory signaling (Cani, Amar et al. 2007, Ghosh, Wang et al. 2020, Jeerawattanawart, Hansakon et al. 2023). Since lower BA and increased 2-methyl-butyrate signify a transition towards proteolytic dysbiosis, the combination of hypoadiponectinemia and SCFA imbalance is anticipated to intensify LPS translocation, oxidative stress, and inflammatory lipid signaling, hence strengthening the NIMETOX-driven metabolic profile seen in severe MDD.

It is important to acknowledge that additional upstream pathways not investigated in our study may mechanistically contribute to the metabolic disorders observed in MDD. In MDD, both catecholamines and cortisol levels are elevated (Maes, Meltzer et al. 1993, Coryell, Young et al. 2006). Catecholamines serve as rapid initiators of lipotoxic stress, as β-adrenergic stimulation activates hormone-sensitive lipase and adipose triglyceride lipase, leading to an immediate increase in circulating non-esterified fatty acids (NEFA), particularly saturated and ω-6 species (Arner 2005, Lafontan and Langin 2009). These fatty acids are preferentially transported to the liver and other tissues; this excess fatty acid flux promotes hepatic DAG accumulation and de novo ceramide synthesis (Summers 2018). Chronic cortisol excess synergistically sustains this condition by promoting lipolysis, increasing hepatic VLDL secretion, and favoring a more atherogenic lipid profile (Djurhuus, Gravholt et al. 2002, Peckett, Wright et al. 2011). Stress-induced endothelial dysfunction and microvascular ischemia further link the HPA-axis and sympathetic activation with enhanced PL remodeling, oxidative vulnerability, and lipotoxic vulnerability (Ghiadoni, Donald et al. 2000, Tyurina, St Croix et al. 2019).

The aforementioned pathways also clarify the substantial influence of ACEs on the metabolic module of the MDD phenotype. Therefore, it has previously been demonstrated that ACEs sensitize or activate SAS, HPA-axis activity, IRS/CIRS, API responses, lipid profiles, increased oxidative stress, reduced antioxidant defenses, and gastrointestinal dysbiosis (Maes, Almulla et al. 2025).

### Consequences of the metabolic aberrations for brain functions

Peripheral metabolomic dysregulation can influence the brain via blood–brain barrier destabilization, endothelial activation, and the transmission of inflammatory lipid signals, thereby contributing to neuroimmune responses and synaptic impairment (Hawkins and Davis 2005, Abbott, Patabendige et al. 2010, Karki and Birukov 2018). Arachidonic acid-enriched phospholipids and LOX-derived products (e.g., leukotrienes) suggest increased bioactive lipid activity within the neurovascular unit (Black and Hoff 1985, Funk 2001). In parallel, the DAG–PKC axis can convert peripheral membrane stress into disruption of the endothelial barrier, as PKC activation promotes tight junction remodeling through the redistribution of occludin, claudin-5, and ZO-1, thereby increasing BBB paracellular permeability, especially under conditions of cytokine stress (Fleegal, Hom et al. 2005, Stamatovic, Dimitrijevic et al. 2006, Willis, Meske et al. 2010, Peng, He et al. 2011). Furthermore, as previously reviewed, all other NIMETOX pathways contribute to the disruption of BBB (Maes, Almulla et al. 2025). The accumulation of DAGs inside neuronal and vascular membranes enhances DAG–PKC signaling, which modulates synaptic effectiveness and may induce circuit-level dysfunction when persistently activated (Lee, Kim et al. 2016). Oxidized phospholipids (OxPLs) function as barrier-active damage-associated molecular patterns (DAMPs), promoting endothelial activation and permeability programs while maintaining inflammatory tone at the neurovascular unit (Fu and Birukov 2009). Phosphatidylserines enriched with polyunsaturated fatty acids serve as redox-sensitive reservoirs for the release of polyunsaturated fatty acids regulated by PLA and for lipoxygenase eicosanoid signaling (Leslie 2015, Kagan, Mao et al. 2017). Simultaneously, oxidative stress-induced phosphatidylserine externalization and release of phosphatidylserine-positive microparticles exacerbate microvascular damage and disrupt neurovascular coupling, therefore affecting cerebral circuitry (Bevers and Williamson 2016). Eoxin C4 is a powerful proinflammatory mediator that can exacerbate perivascular inflammatory signaling in the context of a compromised BBB (Feltenmark, Gautam et al. 2008).

In summary, when the BBB or endothelium is disrupted, lipid mediators including free cholesterol, lipotoxic substances, and leukotrienes, in conjunction with subsequent cytokine cascades and activated M1 and T cells, can sensitize microglia, hinder neuroprotection, and diminish neuroplasticity in hippocampal–amygdalo–prefrontal cortex circuits, correlating with phenotypes seen in MDD (Maes, Almulla et al. 2025).

## Limitations

Notwithstanding meticulous data curation and the removal of non-endogenous metabolites, untargeted metabolomics remains vulnerable to batch effects and platform-specific detection biases. The design is cross-sectional, thereby limiting the ability to establish firm causality. The sample size of 165 participants may be considered to be relatively modest for high-dimensional omics research; the post hoc power for the primary outcome data (regression analysis with OSOD as the final output) was one. Notwithstanding the statistical adjustment for potential confounders (age, sex, BMI, metabolic syndrome, and medication status of the patients), unmeasured variables such as diet, circadian rhythms, and physical activity may have impacted the metabolic variability.

## Conclusions

The principal biomarkers of MDD and its phenotypes include DAG/PKC lipotoxicity, PL remodeling, leukotriene synthesis, fatty acid storage and signaling, mitochondrial redox dysfunction, diminished antioxidant defenses—characterized by reduced PON1 activity, ApoA1, and RCT—alongside the acute phase inflammatory response, gut-derived short-chain fatty acids, and energy homeostatic hormones and adipokines. This NIMETOX profile differentiates MDD from controls with a cross-validated accuracy of more than 95%. PLS-SEM indicates that disturbances in lipotoxicity, PL remodeling, FA storage, overall severity of depression, physiosomatic symptoms, ROI, and present suicidal behaviors are manifestations of a singular underlying latent construct, specifically the “metabolic-phenome of MDD.” It can be inferred that MDD is a metabolic illness. The metabolomic disorders are caused by an imbalance between protective SCFAs and more harmful SCFAs, alterations in energy homeostatic hormones and adipokines, diminished antioxidant defenses, including reduced RCT and PON1, oxidized HDL, the acute phase inflammatory response. These pathways act as intermediaries for the impact of ACEs on the metabolic phenome of MDD. Metabolomic aberrations should be seen as the upstream biological pathway by which many immunological, metabolic, and redox abnormalities manifest as depression symptom severity, recurrence of illness, and suicidality.

## Supporting information

Electronic Supplementary File

## Data Availability

The database compiled during this study will be made available by the corresponding author (MM) upon a reasonable request, following the authors' comprehensive utilization of the data set.

## Ethics approval

This study received approval from the ethics committee of Sichuan Provincial People’s Hospital [Ethics (Research) 2024-203] and was conducted in precise accordance with ethical standards and privacy regulations.

## Consent to participate

Prior to participating in this investigation, each participant provided written informed consent.

## Consent for publication

All authors have provided their consent for the publication of this paper.

## Author’s contributions

Michael Maes: supervision, conceptualization, formal analysis, writing - review and editing. Mengqi Niu: visualization, writing, and recruiting participants. Ping Wang and Annabel Maes: Data analysis. Chenkai Yangyang, Xiaoman Zhuang, Yiping Luo, and Jing Li: recruiting participants. Abbas F Almulla: editing, visualization; Yingqian Zhang: conceptualization, visualization, writing, review, editing.

