## Supplementary material for "NIMETOX-informed Precision Nomothetic Models of Major Depressive Disorder: Group, Phenome, and Individual Signatures": Electronic Supplementary File

**ELECTRONIC SUPPLEMENTARY FILE (ESF)**

**ESF, Assays.**

Preprocessing

Samples were thawed on ice and vortexed for 10 s. 50 μL of sample and 300 μL of extraction solution (ACN: Methanol = 1:4, V/V) containing internal standards were added into a 2 mL microcentrifuge tube. The samples were vortexed for 3 min and then centrifuged at 12000 rpm for 10 min (4 °C). 200 μL of the supernatant was collected and placed at -20 °C for 30 min and then centrifuged at 12000 rpm for 3 min (4 °C). A 180 μL aliquots of supernatant were transferred for LC-MS analysis. Quality control (QC) was obtained by sucking 50μL of the supernatant of each biological sample and vortexing it in a centrifuge tube at room temperature for 60 seconds. Data acquisition and analysis of the experiment were performed at Beijing Junfeix Technology Co., Ltd. The LC-MS analyses were performed using a UPLC system (Vanquish, Thermo Scientific) coupled to an electrospray ionization quadrupole-Orbitrap hybrid high-resolution mass spectrometer (Q Exactive HF-X, Thermo Scientific).

LC-MS

All samples were examined using liquid chromatography (LC) and mass spectrometry (MS) methods. The LC-MS analyses were performed using a UPLC system (Vanquish, Thermo Scientific) coupled to an electrospray ionization quadrupole-Orbitrap hybrid high-resolution mass spectrometer (Q Exactive HF-X, Thermo Scientific). One aliquot was analyzed using positive ion conditions and was eluted from T3 column (Waters ACQUITY Premier HSS T3 Column 1.8 µm, 2.1 mm * 100 mm) using 0.1 % formic acid in water as solvent A and 0.1 % formic acid in acetonitrile as solvent B in the following gradient: 5 to 20 % in 1 min, increased to 99 % in the following 2 mins and held for 1.5 min, then come back to 5 % mobile phase B within 0.1 min, held for 1.4 min. The analytical conditions were as follows, column temperature: 40 °C; flow rate, 0.4 mL/min; injection volume: 4 μL. Another aliquot used negative ion conditions and was the same as the elution gradient of positive mode. All the methods alternated between full scan MS and data dependent MS-n scans using dynamic exclusion. MS analyses were carried out using electrospray ionization in the positive ion mode and negative ion mode using full scan analysis over m/z 75-1000 at 35000 resolutions. Additional MS settings are: ion spray voltage, 3.5 KV or 3.2 KV in positive or negative modes, respectively; Sheath gas (Arb), 30; Aux gas, 5; Ion transfer tube temperature, 320 °C; Vaporizer temperature, 300 °C; Collision energy, 30,40,50 V; Signal Intensity Threshold, 1.00E+06 cps; Top N vs Top speed, 10; Exclusion duration, 3s.

Data processing

The acquired MS data pretreatments including peak picking, peak grouping, retention time correction, second peak grouping, and annotation of isotopes and adducts were performed using XCMS software. LC−MS raw data files were converted into mzXML format and then processed by the XCMS, CAMERA and metaX toolbox implemented with the R software. Each ion was identified by combining retention time (RT) and m/z data. Intensities of each peak were recorded and a three-dimensional matrix containing arbitrarily assigned peak indices (retention time-m/z pairs), sample names (observations) and ion intensity information (variables) was generated. The online KEGG, HMDB database was used to annotate the metabolites by matching the exact molecular mass data (m/z) of samples with those from database. If a mass difference between observed and the database value was less than 10 ppm, the metabolite would be annotated, and the molecular formula of metabolites would further be identified and validated by the isotopic distribution measurements. We also used an in-house fragment spectrum library of metabolites to validate metabolite identification.

**ESF, Statistics**

Regression.

The relationships among the metabolic modules and other NIMETOX indicators were depicted using heatmaps based on Pearson's correlation coefficients. Furthermore, we performed multiple regression analysis to determine the primary biomarkers that predict the metabolic modules. In addition to the manual multiple regression approach, we employed automated regression methods to identify the NIMETOX biomarkers that function as predictors for the metabolic modules. This investigation conducted a ridge regression analysis with a regularization parameter of λ = 0.1 and a tolerance level of 0.4, utilizing Statistica, Windows version 14. Lasso regression was conducted utilizing the CATREG module within SPSS 30. Furthermore, we employed forward stepwise automatic linear modeling analyses, employing an overfitting criterion for the inclusion and exclusion of variables, with a maximum effects threshold set at 5 (using SPSS for Windows, version 30). We conducted a best subset analysis using a criterion to reduce overfitting with SPSS version 30. Following these assessments, we executed a manual regression analysis utilizing SPSS 30 and conducted an exhaustive review of the model statistics, including the F statistic, degrees of freedom, p-values, and the total variance explained by R² (model effect size). We computed the standardized beta coefficients for each predictor, along with their corresponding t statistics and exact p-values. The final models were assessed for multivariate normality, encompassing the residual distributions and P-P plots. An assessment of the variance inflation factor and tolerance was conducted to identify potential issues related to collinearity or multicollinearity. The evaluation of heteroskedasticity was performed using the White test and the modified Breusch-Pagan test to determine homoscedasticity. Furthermore, we performed partial regression analyses on phenome data related to metabolites. The specified regression analyses were performed using IBM SPSS for Windows, version 30, and Statistica, version 14.0. The significance threshold for all statistical analyses was established at 0.05, employing two-tailed tests.

Group profiles

We utilized PLS-DA, OPLS-DA, Random Forest, and SVM to examine the multivariate distinctions between individuals with MDD and control subjects. The first three strategies were employed to evaluate the significance of the biomarkers in this discrimination. OPLS-DA was employed alongside PLS-DA to differentiate class-predictive variance from orthogonal (non-discriminatory) variation in the data. This method produces clearer group differentiation by eliminating structured noise irrelevant to the outcome. In our cross-validation methodologies, all procedures related to feature selection, model training, hyperparameter tuning, and interpretability were conducted solely within the training set, with the testing sample remaining entirely designated for final assessment. This methodology diminished leakage across all analytical levels, guaranteeing unbiased biomarker identification and reliable predictive modeling. The complete sample was arbitrarily divided into training (60%) and testing (40%) subsets. The preliminary biomarker evaluation was confined to the training cohort. All data within the training samples were scaled using Pareto or Z-score transformations. The data were further validated in the testing sample through the application of PLS-DA and SVM.

Our method prevented leakage and ensured independence between exploratory multivariate modeling (training) and subsequent biomarker evaluation (testing). Within the training subset, all biomarkers were analyzed using univariate ANOVAs to identify differences between the MDD and control groups, with adjustments made for potential confounders such as age, sex, BMI, and metabolic syndrome. The key differential biomarkers were identified based on three criteria: a p-value less than 0.05 from ANOVA tests and a VIP score greater than 1 from PLS-DA analysis. The list of candidates was further reduced through iterative refinement of the biomarkers.

The performance of the PLS-DA model across all NIMETOX biomarker matrices was evaluated using R²Y (explained variance in group classification) and Q² (predictive accuracy assessed through cross-validation). To determine the variables that most significantly impact this multivariate structure, variable importance in projection (VIP) scores were computed, and the analysis was reiterated utilizing the highest-ranked VIP features to illustrate their combined effect on PLS-DA discrimination.

A SVM classifier was employed in conjunction with a 10-fold cross-validation approach to assess the robustness and generalizability of the projected accuracy across various resampling folds. SVM identifies an optimal separating hyperplane, whether linear or kernel-based, emphasizing margin maximization over covariance structure. This differs from PLS-DA, which develops latent components designed to maximize covariance between predictors and class membership. The integration of SVM with PLS-DA is synergistic, as SVM provides boundary-based, cross-validated classification performance, while PLS-DA offers interpretable latent structures and variable loadings.

A random forest approach was employed to distinguish MDD from control subjects, utilizing separate training and testing datasets to ensure out-of-sample validation of the classification performance. Random forest detects complex non-linear relationships and higher-order interactions among biomarkers that linear methods fail to adequately capture.

Variable contribution scores were examined to assess the importance of each biomarker, resulting in an internal classification of their predictive significance. This approach profoundly differs from SVM, which optimizes a separation margin, and from PLS-DA, which derives latent linear components that maximize covariance with group membership. PLS-DA emphasizes the overall covariance structure, SVM focuses on boundary optimization, and RF is driven by interactions and hierarchical relationships. The combination of these three complementary methodologies improves robustness, reduces model-specific bias, and increases confidence in the identified biomarker signature.

Phenome profiles

To identify the primary biomarkers associated with the phenome profiles (OSOD, current SI, ROI, and physiosomatic symptoms), we performed PLS-regression and multiple regression analysis following lasso CATREG regression. We utilized PLS-regression to analyze the prediction of phenome scores across the combined training and testing datasets. The model's accuracy was assessed through the use of R²Y and Q² values for all derived latent vectors. PLS-regression was utilized to model OSOD, physiosomatic symptoms, suicidal ideation, and ROI measurements, as continuous outcomes regressed on the NIMETOX biomarker set. The impact of individual biomarkers was evaluated using VIP scores, indicating their relative contribution to the phenome. Consequently, PLS-regression provides a clear multivariate framework that links biological pathways to the clinical presentation of MDD. Additionally, we employed both automated and manual multiple regression analyses to identify the principal biomarkers that predict the phenome scores (see above).

Depression modelling

Partial least squares structural equation modeling (PLS SEM) was utilized to investigate the causal relationships among the NIMETOX biomarkers, the metabolomics modules, and the clinical outcome. The latter was identified as a factor originating from OSOD, psychosomatic symptoms, ROI, and current SI, designated as the "clinical phenome". In addition, we created a latent vector that represents metabolic disorders by integrating the established metabolomics modules. We performed PLS analysis with 5,000 bootstrap samples exclusively when both the outer and inner models satisfied the predetermined quality standards: a) all outer loadings exceeded 0.66 with p < 0.001; b) the latent constructs demonstrated strong construct and convergent validity, as evidenced by an average variance extracted (AVE) greater than 0.5 and composite reliability exceeding 0.8; c) Confirmatory Tetrad Analysis (CTA) verified the model's specification as reflective; and d) discriminant validity was confirmed through the heterotrait-monotrait (HTMT) ratio matrix. Once the specified model quality data criteria are met, we conduct a PLS-SEM path analysis using 5,000 bootstrap samples to obtain path coefficients (including associated p-values) and to calculate both specific and total indirect (mediated) effects, as well as the overall total effects. Furthermore, the overall model fit, as evidenced by the standardized root mean square residual (SRMR), is considered acceptable with a value below 0.08. The redundancy of the constructed model was evaluated through cross-validation employing blindfolding techniques.

Personalized profiles

PLS-regression was utilized to derive case-specific contribution scores that measure the degree to which various biomarker profiles influence the differentiation between MDD and control subjects, as well as the prediction of phenome scores. Scores for individual subjects on the derived latent components reflect each case's position along the biomarker–phenome dimensions. The case-specific ratings indicate the extent to which an individual's biomarker profile aligns with the clinical features. Calculating case-specific contribution scores facilitates the interpretation of biomarker effects on an individual basis rather than exclusively at the aggregate level. This enables a personalized approach by identifying biological factors specific to patients that affect symptom severity and risk. In addition, this approach enables the visualization of how specific metabolites affect a participant's increase in phenome scores.

**ESF, Tables**

ESF, Table 1. Methods to analyze the NIMETOX biomarkers.

|  | Assay. **Derived indices** | Method and kit. **Derived index method.** | Equipment. **Index computation** |
| --- | --- | --- | --- |
| 1 | Albumin (Alb) | Immunoturbidimetric assay (DIAYS DIAGNOSTIC SYSTEM (SHANGHAI) CO., LTD). Sensitivity 0.03 g/L, intra-assay, and inter-assay analytical coefficients of variation (CVs) of 1.96% and 0.67%, respectively. | Fully automated biochemical analyzer (ADVIA 2400, Siemens Healthcare Diagnostic Inc) |
| 2 | Transferrin (Tf) | Bromocresol green method (Beijing Strong Biotechnologies, Inc.), with the intra-assay and inter-assay analytical CVs of 1.20% and 2.10%, respectively. | Fully automated biochemical analyzer (ADVIA 2400, Siemens Healthcare Diagnostic Inc) |
| 3 | Monomeric C-reactive protein (mCRP) | ELISA (BioVendor, Brno, Czech Republic). The sensitivity is 0.63 ng/mL, and the intra-assay and inter-assay CV is < 10% and < 15%, respectively. | Microplate reader (Thermofisher, Sky-High) |
|  | **API index** | **Acute phase inflammatory index:** | **z Alb + zTf + z mCRP** |
| 4 | Total cholesterol (TC) | Cholesterol oxidase-peroxidase-aminoantipyrine-phenol (CHOD-PAP) method (Beijing Strong Biotechnologies, Inc.) with the intra-assay and inter-assay CVs of < 4% and < 6%, respectively. | Fully automated biochemical analyzer (ADVIA 2400, Siemens Healthcare Diagnostic Inc) |
| 5 | Free cholesterol (FC) | CHOD-PAP method (mlbio, China) with the intra-assay and inter-assay CVs of < 3% and < 5%, respectively. | Fully automated biochemical analyzer (ADVIA 2400, Siemens Healthcare Diagnostic Inc) |
|  | **Cholesterol esterification** | **Index of Lecithin-cholesterol acyltransferase (LCAT) activity** | **(1-FC/TC) × 100** |
| 6 | High-density lipoprotein cholesterol (HDL) | Direct method-select inhibition method (Beijing Strong Biotechnologies, Inc.) with the intra-assay and inter-assay CVs of < 4% and < 10%, respectively. | Fully automated biochemical analyzer (ADVIA 2400, Siemens Healthcare Diagnostic Inc) |
| 7 | Apolipoprotein A1 (ApoA1) | Immunoturbidimetric method (Beijing Strong Biotechnologies, Inc.) with the intra-assay and inter-assay CVs of < 3% and < 10%, respectively. | Fully automated biochemical analyzer (ADVIA 2400, Siemens Healthcare Diagnostic Inc) |
| 8 | ApoB | Immunoturbidimetric method (Beijing Strong Biotechnologies, Inc.) with the intra-assay and inter-assay CVs of < 3% and < 10%, respectively. | Fully automated biochemical analyzer (ADVIA 2400, Siemens Healthcare Diagnostic Inc) |
|  | **ApoB/ApoA1** | **Atherogenicity index** | **Ratio ApoB/ApoA1** |
| 9 | ApoE | Immunoturbidimetric method (Beijing Strong Biotechnologies, Inc.) with the intra-assay and inter-assay CVs of < 3% and < 10%, respectively. | Fully automated biochemical analyzer (ADVIA 2400, Siemens Healthcare Diagnostic Inc) |
| 10 | Paraoxonase (PON)1 (Chloromethyl)phenyl acetate CMPAase | Serum CAMPAase activity was quantified by monitoring the hydrolysis of 4- (CMPA, CAS No.: 39720-27-9, Merck, USA) at 280 nm(Brinholi et al. 2024; Maes et al. 2022). The reactions were conducted in UV-transparent 96-well microplates at 25℃, with kinetic absorbance recorded for 4 minutes (16 measurements at 15-second intervals). CMPAase activity was calculated in U/mL based on the molar extinction coefficients of 1.30 mmol/L·cm-1. | Microplate reader (Thermofisher, Sky-High) |
|  | **RCT index** | **Index of reverse cholesterol transport** | **Z composite PON1 + HDL + ApoA1 + LCAT** |
| 11 | Total antioxidant capacity (TOAC) | Colorimetric assay kits (Elabscience, E-BC-K136-M, Wuhan, China). The sensitivity is 0.62 U/mL. The intra-assay and inter-assay CVs are 4.8% and 5.6%, respectively | Microplate reader (Thermofisher, Sky-High) |
|  | **ANTIOX** | **Index of antioxidant capacity** | **Z composite PON1 + HDL + Albumin + TAOC + ApoA1** |
| 12 | Oxidized HDL (oxHDL) | ELISA (Fine Test, EH4858, Wuhan, China). The sensitivity is 0.938 ng/mL, and the intra-assay and inter-assay CVs of 5.12% and 5.22%, respectively | Microplate reader (Thermofisher, Sky-High) |
| 13 | Oxidized LDL(oxLDL) | ELISA (Elabscience, E-EL-H6021, Wuhan, China). The sensitivity is 37.5 pg/mL, and the intra-assay and inter-assay CVs of 6.62% and 7.04%, respectively. | Microplate reader (Thermofisher, Sky-High) |
|  | **HDL/oxHDL** | Index of increased oxidation vulnerability of HDL particles | **Z composite: HDL - oxHDL** |
|  | **oxHDL/oxLDL** | Index of oxidized HDL versus LDL | **Z composite: oxHDL - oxLDL** |
| 14 | Insulin (INS) | Atellica IM insulin assay kit (direct chemiluminescence method, Siemens Healthcare Diagnostics Inc.) The intra-assay and inter-assay analytical CVs are 1.8% and 3.6%, respectively. | Fully automated chemiluminescence immunoassay analyzer (Siemens Healthcare Diagnostics Inc.). |
| 15 | Fasting blood glucose (FBG) | Glucose assay kit (glucose oxidase method, Gcell, Beijing Strong Biotechnologies, Inc. intra-assay and inter-assay analytical coefficients of variation (CVs) of 3.2% and 6.8% respectively | Fully automated biochemical analyzer (ADVIA 2400, Siemens Healthcare Diagnostics Inc.) |
|  | **IR index** | **Insulin resistance (IR) index** | **Z composite: Insulin + Glucose** |
| 16 | Cytokines, chemokines, growth factors | Luminex xMAP technology using Bio-Rad 48 -Human Cytokines, Chemokines and Growth Factor Assays. Intra-assay < 5.0%. | Luminex 200 system |
|  | **M1 macrophage** | **Index of activated macrophage M1 phenotype** | **Z composite: IL-1α, IL-1β, sIL-1RA, TNF-α, IL-6** |
|  | **Thelper-1 (Th1)** | **Index of activated Th2 phenotype** | **Z composite: IFN-α2, IFN-γ, IL-2, IL-12p70** |
|  | **Th17** | **Index of activated Th17 phenotype IL-6, IL-17A** | **Z composite: IL-6, IL-17** |
|  | **IRS** | **Index of activation of the immune-inflammatory responses system** | **Z compositeIL-1α, IL-1β, TNF-α, IL-6, IFN-α2, IFN-γ，IL-2, IL-12p70, IL-17** |
|  | **Th2** | **Index of activated Th17 phenotype** | **Z composite: IL-4, IL-5, IL-10** |
|  | **CIRS** | **Index of the compensatory immunoregulatory system** | **Z composite: IL-4, IL-5, IL-10, sIL-1RA** |
|  | **TNF signaling** | **Index of increased tumor necrosis factor (TNF) signaling** | **Z composite: TNF-α, TNF-β, TRAIL** |
|  | EGF | Epidermal growth factor (EGF). Bio-Rad 48 -Human Cytokines, Chemokines and Growth Factor Assays | Luminex 200 system |
|  | sCD40L | Soluble CD40L molecule (sCD40L). Bio-Rad 48 -Human Cytokines, Chemokines and Growth Factor Assays | Luminex 200 system |
|  | Fms-like tyrosine kinase 3 ligand (Flt3L) | Bio-Rad 48 -Human Cytokines, Chemokines and Growth Factor Assays | Luminex 200 system |
| 17 | Metabolic hormones: Ghrelin, gastric inhibitory polypeptide (GIP), glucagon-like peptide (GLP‑1), Glucagon, Leptin, Secretin | Multiplex magnetic bead‑based immunoassay using Human Metabolic Hormone Panel V3. | Luminex 200 system |
| 18 | Adipokines: Resistin, Plasminogen activator inhibitor (PAI)‑1, Adiponectin | Multiplex magnetic bead‑based immunoassay using the Human Adipokine Magnetic Bead Panel 1 | Luminex 200 system |
|  | **GAP index** | **Ghrelin adiponectin PAI-1 index** | **Z composite (all three positive sign)** |
| 19 | Short chain fatty acids (SCFAs) | The assay of Butyrate (BA), acetate (AA), propionate (PA), succinate (SA), isovalerate (isoVA), 2-methy-butyrate (MBA), hexanoic acid (HexaA), pentanoic (valeric) acid (PA), isobutyrate (isobar) was performed using liquid chromatography-mass spectrometry (LC-MS) at Beijing Junfeix Technology Co., Ltd. | ACQUITY TA-S Triple Quadrupole Tandem Mass-Spectrometer (Waters, USA) |
|  | **Protective SCFA** | **Butyrate, acetate, propionate** | **Z composite score (all three positive sign)** |

ESF, Table 2. Demographic, clinical and metabolomics modules scores in inpatients with major depressive disorder (MDD) and healthy controls (HCs).

| **Variables** | **HC (n = 40)** | **MDD (n = 125)** | **F/χ^2^** | **df** | ***p*** |
| --- | --- | --- | --- | --- | --- |
| Age (years) | 37.1 (13.8) | 35.7 (12.1) | 0.37 | 1/163 | 0.542 |
| Female / Male | 27 / 13 | 87 / 38 | 0.06 | 1 | 0.802 |
| Marriage status (yes / no) | 17 / 23 | 69 / 56 | 1.96 | 1 | 0.162 |
| Living status (Uban / rural) | 36 / 4 | 114 / 11 | FET | - | 0.760 |
| Education (years) | 13.9 (4.3) | 13.5 (3.3) | 0.26 | 1/163 | 0.613 |
| Metabolic syndrome (yes/no) | 10 / 29 | 22 / 102 | 1.17 | 1 | 0.279 |
| Body mass index (kg/m^2^) | 23.52 (4.07) | 22.30 (3.36) | 3.58 | 1/163 | 0.060 |
| Smoking (yes / no) | 3 / 37 | 24 / 101 | FET | - | 0.091 |
| Overall severity of depression (z score) | -1.464 (0.671) | 0.537 (0.510) | MWUT | - | <0.001 |
| Physiosomatic symptoms (z score) | -1.300 (0.421) | 0.477 (0.671) | MWUT | - | <0.001 |
| Current suicidal ideation (z score) | -0.774 (0.262) | 0.269 (1.021) | MWUT | - | <0.001 |
| Recurrence of illness (ROI) (z score) | -1.057 (0.150) | 0.367 (0.903) | MWUT | - | <0.001 |
| Lipotoxicity ( z core) | -1.351 (0.101) | 0.408 (0.056) | 230.34 | 1/154 | <0.001 |
| PL remodeling (z score) | -1.504 (0.086) | 0.467 (0.048) | 393.70 | 1/154 | <0.001 |
| Ether Lipids (z score) | 0.879 (0.143) | -0.273 (0.080) | 49.26 | 1/154 | <0.001 |
| Mito+ATP+Redox (z score) | -0.934 (0.139) | 0.298 (0.077) | 59.49 | 1/154 | <0.001 |
| Retinoid-detox | 0.454 (0.158) | -0.140 (0.088) | 10.71 | 1/154 | <0.001 |
| Fatty acid storage/modelling (z score) | -1.189 (0.120) | 0.366 (0.067) | 126.36 | 1/154 | <0.001 |
| G-metabolomics (z score) | -1.536 0.083 | 0.478 (0.046) | 446.41 | 1/154 | <0.001 |

Data are shown as mean ±SD (results of analysis of variance), except the metabolomics data which are shown as marginal estimated mean values ±SE (results of GLM analysis after covarying for age, sex, metabolic syndrome and body mass index), and frequencies (results of chi-square tests). FET: Fisher’s exact probability test; MWUT: Mann-Whitney U test.

ESF, Table 3. Specific indirect effects (PLS model #1).

| **Paths** | **t** | ***p*** |
| --- | --- | --- |
| **AA -> Metabolism -> Phenome** | **-2.51** | **0.012** |
| **ACEs -> Metabolism -> Phenome** | **3.889** | **<0.001** |
| **GAPindex -> Metabolism -> Phenome** | **-3.722** | **<0.001** |
| **ACEs -> GAPindex -> Metabolism** | **2.397** | **0.017** |
| **HDLparticle -ANTIOX -> Metabolism -> Phenome** | **-4.083** | **<0.001** |
| **API response -> GAPindex -> Metabolism** | **2.638** | **0.008** |
| **MBA -> Metabolism -> Phenome** | **2.858** | **0.004** |
| Sex -> ACEs -> AA | 1.897 | 0.058 |
| **ACEs -> GAPindex -> Metabolism -> Phenome** | **2.275** | **0.023** |
| **Sex -> ACEs -> GAPindex** | **-2.015** | **0.044** |
| **SexAbuse -> Metabolism -> Phenome** | **2.189** | **0.029** |
| ACEs -> HDLparticle -ANTIOX -> Metabolism -> Phenome | 1.807 | 0.071 |
| Sex -> ACEs -> HDLparticle -ANTIOX | 1.846 | 0.065 |
| Sex -> ACEs -> HDLparticle -ANTIOX -> Metabolism | -1.761 | 0.078 |
| **Sex -> ACEs -> Metabolism** | **-2.545** | **0.011** |
| ACEs -> AA -> Metabolism -> Phenome | 1.81 | 0.07 |
| Sex -> ACEs -> HDLparticle -ANTIOX -> Phenome | -1.333 | 0.183 |
| **Sex -> ACEs -> Phenome** | **-2.733** | **0.006** |
| Sex -> SexAbuse -> Metabolism -> Phenome | -1.454 | 0.146 |
| Immune activation -> API response -> GAPindex -> Metabolism | 1.707 | 0.088 |
| Sex -> ACEs -> HDLparticle -ANTIOX -> Metabolism -> Phenome | -1.746 | 0.081 |
| Sex -> ACEs -> AA -> Metabolism -> Phenome | -1.506 | 0.132 |
| Sex -> ACEs -> GAPindex -> Metabolism -> Phenome | -1.78 | 0.075 |
| ACEs -> HDLparticle -ANTIOX -> Metabolism | 1.794 | 0.073 |
| ACEs -> HDLparticle -ANTIOX -> Phenome | 1.384 | 0.167 |
| **API response -> HDLparticle -ANTIOX -> Metabolism** | **3.625** | **<0.001** |
| **API response -> HDLparticle -ANTIOX -> Phenome** | **2.22** | **0.026** |
| **Immune activation -> API response -> GAPindex** | **-2.098** | **0.036** |
| Sex -> SexAbuse -> Metabolism | -1.478 | 0.139 |
| **Immune activation -> API response -> HDLparticle -ANTIOX** | **-2.303** | **0.021** |
| ACEs -> AA -> Metabolism | 1.827 | 0.068 |
| ACEs -> AA -> Phenome | 1.541 | 0.123 |
| **Sex -> ACEs -> Metabolism -> Phenome** | **-2.482** | **0.013** |
| **API response -> HDLparticle -ANTIOX -> Metabolism -> Phenome** | **3.175** | **0.002** |
| **API response -> GAPindex -> Metabolism -> Phenome** | **2.449** | **0.014** |
| Sex -> ACEs -> GAPindex -> Metabolism | -1.874 | 0.061 |
| **Immune activation -> API response -> HDLparticle -ANTIOX -> Metabolism** | **2.009** | **0.045** |
| Immune activation -> API response -> HDLparticle -ANTIOX -> Phenome | 1.54 | 0.124 |
| Sex -> ACEs -> AA -> Metabolism | -1.535 | 0.125 |
| Sex -> ACEs -> AA -> Phenome | -1.485 | 0.138 |
| Immune activation -> API response -> GAPindex -> Metabolism -> Phenome | 1.638 | 0.102 |
| Immune activation -> API response -> HDLparticle -ANTIOX -> Metabolism -> Phenome | 1.891 | 0.059 |

ESF, Table 4. Total indirect effects (PLS model #1).

| **Paths** | **t** | ***p*** |
| --- | --- | --- |
| **AA -> Phenome** | **-2.51** | **0.012** |
| **ACEs -> Metabolism** | **3.438** | **0.001** |
| **ACEs -> Phenome** | **5.995** | **<0.001** |
| **API response -> Metabolism** | **4.406** | **<0.001** |
| **API response -> Phenome** | **4.638** | **<0.001** |
| **GAPindex -> Phenome** | **-3.722** | **<0.001** |
| **HDLparticle -ANTIOX -> Phenome** | **-4.083** | **<0.001** |
| **Immune activation -> GAPindex** | **-2.098** | **0.036** |
| **Immune activation -> HDLparticle -ANTIOX** | **-2.303** | **0.021** |
| **Immune activation -> Metabolism** | **2.145** | **0.032** |
| **Immune activation -> Phenome** | **2.166** | **0.03** |
| **MBA -> Phenome** | **2.858** | **0.004** |
| Sex -> AA | 1.897 | 0.058 |
| **Sex -> GAPindex** | **2.015** | **0.044** |
| Sex -> HDLparticle -ANTIOX | 1.846 | 0.065 |
| **Sex -> Metabolism** | **-3.209** | **0.001** |
| **Sex -> Phenome** | **-3.386** | **0.001** |
| **SexAbuse -> Phenome** | **2.189** | **0.029** |

ESF, Table 5. Total effects (PLS model#1).

| **Paths** | **t** | ***p*** |
| --- | --- | --- |
| **AA -> Metabolism** | **-2.548** | **0.011** |
| **AA -> Phenome** | **-3.493** | **<0.001** |
| **ACEs -> AA** | **-2.321** | **0.02** |
| **ACEs -> GAPindex** | **-2.671** | **0.008** |
| **ACEs -> HDLparticle -ANTIOX** | -1.955 | 0.051 |
| **ACEs -> Metabolism** | **6.126** | **<0.001** |
| **ACEs -> Phenome** | **7.944** | **<0.001** |
| **API response -> GAPindex** | **-3.887** | **<0.001** |
| **API response -> HDLparticle -ANTIOX** | **-5.571** | **<0.001** |
| **API response -> Metabolism** | **4.406** | **<0.001** |
| **API response -> Phenome** | **4.638** | **<0.001** |
| **GAPindex -> Metabolism** | **-4.341** | **<0.001** |
| **GAPindex -> Phenome** | **-3.722** | **<0.001** |
| **HDLparticle -ANTIOX -> Metabolism** | **-4.736** | **<0.001** |
| **HDLparticle -ANTIOX -> Phenome** | **-5.287** | **<0.001** |
| **Immune activation -> API response** | **2.699** | **0.007** |
| **Immune activation -> GAPindex** | **-2.098** | **0.036** |
| **Immune activation -> HDLparticle -ANTIOX** | **-2.303** | **0.021** |
| **Immune activation -> Metabolism** | **2.145** | **0.032** |
| **Immune activation -> Phenome** | **2.166** | **0.03** |
| **MBA -> Metabolism** | **2.972** | **0.003** |
| **MBA -> Phenome** | **2.858** | **0.004** |
| **Metabolism -> Phenome** | **10.088** | **<0.001** |
| Sex -> AA | 1.897 | 0.058 |
| **Sex -> ACEs** | **-3.575** | **<0.001** |
| **Sex -> GAPindex** | **2.015** | **0.044** |
| Sex -> HDLparticle -ANTIOX | 1.846 | 0.065 |
| **Sex -> Metabolism** | **-3.209** | **0.001** |
| **Sex -> Phenome** | **-3.386** | **0.001** |
| **Sex -> SexAbuse** | **-1.977** | **0.048** |
| **SexAbuse -> Metabolism** | **2.292** | **0.022** |
| **SexAbuse -> Phenome** | **2.189** | **0.029** |

ESF, Table 6. Total effects (PLS model #2).

| **Paths** | **t** | ***p*** |
| --- | --- | --- |
| **ACEs -> API response** | **4.230** | **<0.001** |
| **ACEs -> GAPindex** | **-4.002** | **<0.001** |
| **ACEs -> HDLparticle -ANTIOX** | **-3.788** | **<0.001** |
| ACEs -> Immune activation | -0.857 | 0.391 |
| **ACEs -> Metabolic _phenome** | **8.488** | **<0.001** |
| **ACEs -> Protective _SCFAs** | **-2.295** | **0.022** |
| **API response -> HDLparticle -ANTIOX** | **-5.690** | **<0.001** |
| **API response -> Metabolic _phenome** | **4.106** | **<0.001** |
| **GAPindex -> Metabolic _phenome** | **-3.318** | **0.001** |
| **HDLparticle -ANTIOX -> Metabolic _phenome** | **-5.713** | **<0.001** |
| **Immune activation -> API response** | **3.094** | **0.002** |
| **Immune activation -> GAPindex** | **-2.001** | **0.045** |
| **Immune activation -> HDLparticle -ANTIOX** | **-2.503** | **0.012** |
| **Immune activation -> Metabolic _phenome** | **2.507** | **0.012** |
| **MBA -> Metabolic _phenome** | **2.999** | **0.003** |
| **Protective _SCFAs -> Metabolic _phenome** | **-3.489** | **<0.001** |
| **Sex -> ACEs** | **-3.575** | **<0.001** |
| **Sex -> API response** | **-2.562** | **0.010** |
| **Sex -> GAPindex** | **2.481** | **0.013** |
| **Sex -> HDLparticle -ANTIOX** | **2.809** | **0.005** |
| Sex -> Immune activation | 0.811 | 0.418 |
| **Sex -> Metabolic _phenome** | **-3.385** | **0.001** |
| Sex -> Protective _SCFAs | 1.871 | 0.061 |
| **Sex -> SexAbuse** | **-1.977** | **0.048** |
| **SexAbuse -> Metabolic _phenome** | **2.323** | **0.020** |
